# Associations of ^18^F-RO-948 Tau PET with Fluid AD Biomarkers, Centiloid, and Cognition in the Early AD Continuum

**DOI:** 10.1101/2025.07.09.25331011

**Authors:** Mahnaz Shekari, Armand González Escalante, Marta Milà-Alomà, Carles Falcon, David López-Martos, Gonzalo Sánchez-Benavides, Anna Brugulat-Serrat, Aida Niñerola-Baizán, Nicholas J. Ashton, Thomas K. Karikari, Juan Lantero-Rodriguez, Laia Montoliu-Gaya, Anniina Snellman, Theresa A Day, Jeffrey L. Dage, Paula Ortiz-Romero, Matteo Tonietto, Edilio Borroni, Gregory Klein, Gwendlyn Kollmorgen, Margherita Carboni, Clara Quijano-Rubio, Eugeen Vanmechelen, Carolina Minguillón, Karine Fauria, Andrés Perissinotti, José Luis Molinuevo, Henrik Zetterberg, Kaj Blennow, Oriol Grau-Rivera, Marc Suárez-Calvet, Juan Domingo Gispert, ALFA Study

## Abstract

**INTRODUCTION:** We investigated the presence of neurofibrillary tangles (NFT) and their association with amyloid plaques and fluid biomarkers of Alzheimer’s disease (AD) in cognitively unimpaired individuals (CU).

**METHODS:** Ninety-nine CU individuals from the ALFA+ cohort underwent tau PET using ^18^F-RO-948, amyloid PET, MRI, cognitive assessments, and (CSF)/plasma biomarkers. Tau PET SUVRs calculated in Braak regions, and associations with AD biomarkers were assessed. Thresholds for positivity were derived, and ROC analysis evaluated biomarker accuracy in predicting tau PET positivity.

**RESULTS:** Nine cases were Braak-I/II positive, five Braak-III/IV, and one Braak-V/VI. For CSF biomarkers, the strongest associations with Braak-I/II reached r=0.58 [95%CI:0.26-0.79, p<0.001, CSF ptau217] and, for plasma, r=0.25 [95%CI:0.03-0.45, p=0.01, plasma ptau181/Aβ42], but showed low PPVs [0.09-0.33] for Braak-I/II positivity.

**DISCUSSION:** ^18^F-RO-948 PET was capable of detecting positive cases in the earliest preclinical AD stages. The studied fluid biomarkers alone showed limited accuracy for screening CU individuals for tau PET positivity.

## 1 BACKGROUND

Neurofibrillary tangles (NFT), composed of hyperphosphorylated tau, are the defining characteristic of Alzheimer’s Disease (AD), together with fibrillar amyloid-β (Aβ) deposits in the brain^1–4^. Typically, NFT appears later than Aβ in the preclinical stages of the disease and is a strong predictor of imminent neurodegeneration^5^ and cognitive decline. Several radiotracers have shown their capacity to image in vivo the amount and spread of NFT pathology^6^. These tracers have been validated against neuropathology^7^, and some have been or are being developed for diagnostic clinical use^8^. Several tau positron emission tomography (PET) radiotracers, including ¹ F-flortaucipir and ¹ F-MK-6240, have been validated through postmortem studies, showing strong concordance with the distribution and burden of tau pathology in the brain^9^. Furthermore, head-to-head comparisons among ¹ F-flortaucipir, ¹ F-RO-948, and ¹ F-MK-6240 have demonstrated high inter-tracer agreement in tau PET signal patterns^10,11^, reinforcing their reliability in capturing tau pathology in vivo and supporting their validity against neuropathological standards.

The spread of NFT in AD is typically believed to follow neuropathologically defined Braak staging^4,12^. According to the prototypical Braak spread pattern, tau starts accumulating in medial temporal regions (Braak I/II) before spreading to limbic regions (Braak III/IV) and finally to the whole cortical mantle (Braak V/VI). This pattern of cortical spread has been formalized into the hierarchical Braak staging system, which is part of the gold-standard diagnostic workup for AD at autopsy^3^. However, recent in vivo tau PET studies across multiple cohorts suggest that several subtypes of tau spread can be identified with distinct demographic characteristics, cognitive profiles, and longitudinal outcomes^13,14^. This highlights the added value of tau PET for the study of the pathological diversity of AD, and its relationship with the observed heterogeneity in clinical symptoms^15^.

While tau PET imaging can accurately detect and quantify NFT pathology, there is an urgent need for inexpensive and minimally invasive blood AD biomarkers to detect early disease changes. Efficient blood-based biomarkers may also guide treatment decisions and support clinical management in practice. In recent years, several highly specific and potentially useful blood-based biomarkers for AD have been identified^16^, with ptau variants among the most promising. Several studies have shown that ptau181, ptau217, and ptau231 are highly specific for AD and rise early on the preclinical AD continuum^17,18^. However, studies against neuropathology show that plasma ptau181 and ptau217 mark both Aβ and NFT pathology, whereas plasma ptau231 is mainly associated with Aβ^19,20^. Recent reports on cerebrospinal fluid (CSF) biomarkers, particularly CSF ptau20521 and CSF MTBR-tau24322, suggest that they are more specific to aggregated insoluble tau pathology; however, their blood-based counterparts remain to be fully characterized and validated.

In summary, while blood-based biomarkers’ capacity to predict the presence of Aβ pathology using amyloid PET is good^23–25^ their concordance with tau PET remains low. This is further aggravated in the preclinical stages of AD, where changes in NFT are subtle compared to more advanced stages^26^. Therefore, there is a need for blood-based biomarkers that can specifically inform on the presence of tau pathology in preclinical AD. Additionally, there is limited evidence in the literature reporting the extent to which different regional patterns of tau pathology can be detected in preclinical stages of AD.

In this work, we aimed to assess NFT pathology in the early preclinical AD continuum, as assessed by 18F-RO-948 tau PET imaging, and its relationship with other AD biomarkers. To this end, we acquired Aβ and tau PET, quantified several tau phospho-forms in the cerebrospinal fluid and blood, and assessed their agreement in a sample of the ALFA+ cohort of cognitively unimpaired participants selected based on their baseline levels of Aβ load^27^.

## 2 METHODS

### 2.1 Participants

A total of 211 cognitively unimpaired (CU) participants with available amyloid PET scans, T1-weighted magnetic resonance imaging (MRI), fluid AD biomarkers, and cognition scores were selected from the ALFA+ cohort^27^. Of these, a subset of 100 participants underwent tau PET scanning, selected based on CSF-derived AT staging, where ‘A’ indicates amyloid positivity (defined by the CSF Aβ42/40 ratio cut-off of 0.071) and ‘T’ indicates tau positivity (defined by the CSF ptau181 cut-off of 24 pg/ml), aiming for the most balanced distribution across the four groups (A–T–, A+T–, A+T+, and A–T+)^28^. More information about the tau PET study design can be found at https://clinicaltrials.gov/study/NCT04482660. ALFA+ is a longitudinal study of 450 CU individuals enriched for a family history of AD and APOE ε4 carriership. Inclusion and exclusion criteria, along with study details, have been previously described^27^.

All participants gave written informed consent. The ALFA+ study was approved by the independent ethics committee ‘Parc de Salut Mar’, Barcelona, and is registered at Clinicaltrials.gov (Identifier: NCT02485730).

### 2.2 PET and MRI Acquisition

Amyloid PET scans were conducted between March 6, 2020, and June 27, 2022, while tau PET scans were performed between March 29, 2021, and February 7, 2023. Aβ-PET and tau PET data acquisition was performed for 20 min (4 frames × 5min), on a Siemens Biograph mCT scanner, following a cranial computed tomography (CT) scan for attenuation correction. Tau PET images were acquired 70 to 90 min after injecting 368.75±13.02 MBq of ^18^F-RO-948 and Aβ-PET scans were collected 90 to 110 min after injecting ∼185 MBq of ^18^F-flutemetamol. Both tau PET and Aβ-PET scans were reconstructed using an ordered subset expectation maximization (OSEM) algorithm and incorporating time of flight (TOF) and point spread function modeling (Tau PET: 4 iterations, 21 subsets, 4 mm; Aβ-PET: 8 iterations, 21 subsets, 3 mm).

T1-weighted MRIs were obtained using a 3T scanner (Ingenia CX, Philips Healthcare, Best, The Netherlands) with a 32-channel head coil and a 3D Turbo Field Echo sequence with the following parameters: TE/TR=4.6/9.9 ms, Flip Angle = 8°, and voxel size= 0.75×0.75×0.75 mm.

### 2.3 PET Quantification

Prior to preprocessing, all PET scans underwent quality control. One tau PET scan was excluded because it was acquired significantly later than the recommended post-injection time, making the data unreliable. Next, the images were processed with SPM12 (www.fil.ion.ucl.ac.uk/spm). Standardized uptake value ratio (SUVR) parametric images of ^18^F-RO-948 were created in the subject space by dividing the voxel intensities by the average intensity in a reference region covering the inferior cerebellum using the SUIT atlas^29,30^. For creating Braak region of interests (ROIs), a composite atlas was created using the predefined regions of interest defined elsewhere^31^. To create a subject-based Braak atlas, first, the Braak atlas template was moved to the native space using the deformation field, extracted from segmenting the T1-weighted MRI. Next, the Braak atlas template was coregistered and resliced with T1-weighted MRI. Then a binary grey matter (GM) mask was created where the probabilities of GM>WM (white matter) and GM>CSF were assigned to 1, and the rest were assigned to zero. Finally, the Braak atlas template mask was multiplied by the binary GM mask to create a subject-based Braak mask (Figure_S1 (B)). In addition to the subject-based Braak mask, subject-based meninges and skull masks were created by segmenting the corresponding CT scan of the participant (Figure_S1 (A)). Mean meningeal and skull uptakes were created for all participants. Pearson correlation between Braak ROI uptake and meningeal SUVR was calculated for all participants. A statistically significant correlation between any Braak ROI and meningeal uptake was interpreted as meningeal contamination of the cortical Braak ROI. To minimize this contamination, only individuals with meningeal SUVR>2 was selected for cortical sharpening. The meningeal mask was then dilated 1 to 10 times for each participant with meningeal SUVR above 2. Voxels overlapping between the dilated meningeal mask and cortical ROIs were removed to refine the cortical mask. Cortical SUVR for each Braak stage was then calculated using this sharpened cortical mask. The optimal dilation level was determined when SUVR values became independent of further sharpening (Figure_S2 and Figure_S3). As a final quality control step, Pearson correlation between Braak ROIs and meningeal SUVR was recalculated, and the absence of a significant correlation was considered evidence of reliable quantification.

Finally, the SUVR was calculated in cortical regions representing different Braak stages: entorhinal cortex (Braak I/II), limbic (Braak III/IV), and neocortical (Braak V/VI) regions. Regional positivity per Braak stage (Braak I/II+, Braak III/VI+, Braak V/VI+) was calculated as the mean plus two standard deviations of the SUVR in each Braak stage for the reference control group (CSF A-T-individuals). Tau PET staging for each Braak ROI and the global region was determined using the calculated thresholds. In addition to SUVR, all images were quantified using the CenTauR-z method^32^. CenTauR-z was developed to standardize tau PET quantification across different tracers by generating z-scored SUVR values for predefined cortical ROIs. The meta-ROI template used for this analysis is available on the GAAIN website^33^. Further details on the preprocessing steps can be found in the original publication^32^.

Aβ-PET scans were quantified in Centiloid (CL) units, using a validated in-house standard pipeline^34,35^. In brief, an average PET image was created after realigning 4 frames to correct for any possible motions between frames. Then, the PET scans were coregistered with their corresponding T1-weighted MRI and warped to Montreal Neurological Institute (MNI) space. The SUVR maps were created using the predefined GAAIN whole cerebellum mask as the reference region. Finally, SUVR was calculated for the GAAIN pre-defined cortical target region and transformed into the Centiloid^36^.

### 2.4 Plasma and CSF Biomarkers

Fluid biomarker samples were collected within 3.00±4.08 months of the amyloid PET scan and 9.42±9.31 months of tau PET scans. CSF and blood biomarkers were collected and processed using different assays, following standard procedures^37,38^. CSF ptau205, ptau235, and NTA-tau (N-terminal tau), as well as Plasma pTau181, pTau231, and Aβ42 concentrations were measured using either commercial or in-house single-plex assays on the Simoa HD-X platform (Quanterix, Billerica, MA, USA). CSF ptau181, Aβ40, and Aβ42 were measured using the electrochemiluminescence Elecsys immunoassay on a fully automated cobas e601 module (both Roche Diagnostics International Ltd., Rotkreuz, Switzerland). CSF [ptau181, Aβ40, Aβ42, t-tau] and plasma [Aβ40, Aβ42, ptau181] were measured using the Roche NeuroToolKit immunoassays (Roche Diagnostics International Ltd.) on a cobas e411 or e601 analyzer. Additionally, ptau217 in both CSF and plasma was measured using an Eli Lilly assay on the Meso Scale Discovery platform (MSD)^39^. All participants were categorized in AT stages according to CSF, using pre-established CSF-based cut-off values (CSF Aβ42/40 < 0.071 (A+) and CSF ptau181 > 24 pg/mL (T+))^28^.

### 2.5 Neuropsychological evaluation

A modified version of the Preclinical Alzheimer’s Cognitive Composite (PACC) score, consisting of attention, executive, language, memory, and visual composites was available for all participants^40–42^. All raw test scores were standardized into z-scores using the mean and standard deviation (SD) from CU CSF A-T-participants as a reference and then averaged into a composite score^42^.

### 2.6 Statistical Analyses

Descriptive statistics were calculated for the main demographic variables of the participants grouped by the CSF-based AT status. Correlations between Centiloid, ^18^F-RO-948 SUVRs in each Braak region, and AD biomarkers were assessed using partial correlation, adjusted for age, sex, APOE 4 carriership, and the time interval between PET imaging and fluid biomarker collection (ΔTime).

Eq.1 Braak SUVR ∼ 1+ Biomarker + Age + Sex + APOE 4 + ΔTime

Receiver Operating Characteristic (ROC) analyses were performed to assess the ability of fluid biomarkers to predict Braak I/II positivity in CU individuals. Analyses were performed using models that included biomarkers alone as well as models adjusted for age, sex, and APOE 4 carriership. Youden’s Index (YI) was used to determine optimal thresholds using the results of the adjusted model. In particular, the optimal CL cut-off to predict Braak I/II positivity (Centiloid-Braak I/II+) was also calculated. AUC values of [0.7-0.8] were considered as acceptable, [0.8–0.9] as excellent and above 0.9 as outstanding performance in discrimination. Additionally, the positive predictive value (PPV) and negative predictive value (NPV) of each biomarker were calculated.

A posteriori, we defined four PET-based AT stages based on amyloid PET (A) and tau PET (T) positivity^43^. Under this scheme, participants were classified as below:

- PET (A-T-): Centiloid < 12 & Braak I/II < Braak I/II+
- PET (A(Gz)T-): 12 ≤ Centiloid < Centiloid-Braak I/II+ & Braak I/II < Braak I/II+
- PET(A+T-): Centiloid ≥ CentiloidBraak I/II+ & Braak I/II < Braak I/II+
- PET (A+T+): Centiloid ≥ CentiloidBraak I/II+ & Braak I/II ≥ Braak I/II+

On top of the data-driven cut-offs for CL and tau PET positivity, we also applied a CL <12 threshold based on prior literature supporting its sensitivity for detecting early amyloid abnormalities. All biomarkers were z-scored using the CSF-based A-T-group as the reference for standardized comparisons. To examine group differences, we used the Kruskal-Wallis test across the four PET-derived AT groups and applied Bonferroni correction for multiple comparisons.

To assess the magnitude of biomarker changes across PET-defined disease stages, we additionally calculated fold changes using the raw (non-z-scored) fluid biomarker values. Each value was divided by the mean of the corresponding biomarker in the PET A-T-reference group (individuals who were negative for both amyloid and tau PET). These fold changes offer a complementary view of absolute biomarker shifts associated with disease progression. For all analyses, p-values < 0.05 were considered statistically significant.

## 3 RESULTS

Table 1 presents demographic characteristics, Centiloid values, hippocampal volumes, regional tau PET SUVRs across Braak stages, CenTauR-z scores, and PACC scores for participants with available tau and amyloid PET scans after quality control, stratified by CSF-based AT status. Demographic information of 212 participants included in the Centiloid analysis can be found in the supplementary, Table_S1.

**Table 1.** Demographic characteristics, Centiloid values, and PACC scores of participants, stratified by CSF-based amyloid (A) and tau (T) status. Biomarker samples were collected within 9.42 ± 9.31 months of tau PET imaging. Participants were classified into AT stages using pre-established cutoffs21 based on CSF Aβ42/40<0.071 and ptau181>24(pg/ml) measured with the NeuroToolKit, a panel of robust prototype assays and Elecsys® immunoassays (Roche Diagnostics International Ltd). Between-group comparisons were performed using ANOVA, followed by Bonferroni post hoc corrections, with A-T-as the reference group. Statistical significance (p < 0.001) for multiple comparisons is indicated by “1**”.

|  | **Total** | **A-T-** | **A+T-** | **A+T+** | **A-T+** |
| --- | --- | --- | --- | --- | --- |
| **N** | 99 | 27 | 33 | 28 | 9 |
| **Age, Mean (SD) [53-78 years]** | 65.15±5.00 | 64.25±4.97 | 65.12±4.94 | 66.32±5.59 | 64.22±3.03 |
| **Sex, N (%) [Female]** | 57 (57.57%) | 14 (51.85%) | 21 (63.63%) | 18 (64.28%) | 3 (33.33%) |
| **APOE-ε4, N (%) [Carriers]** | 63 (63.63%) | 12 (44.44%) | 26 (78.78%) | 19 (67.85%) | 4 (44.44%) |
| **Years of Education** | 13.35±3.78 | 14.14±3.86 | 14.12±3.48 | 11.92±3.71 | 13.77±3.52 |
| **CSF Aβ42/40, Mean (SD)** | 0.06±0.02 | 0.08±0.01 | 0.05±0.01 | 0.04±0.01 | 0.11±0.02 |
| **CSF ptau181, Mean (SD), pg/mL** | 21.92±9.40 | 15.12±4.47 | 17.41±4.09 | 31.96±8.98** | 27.28±4.74** |
| **CSF ptau181/Aβ40** | (1.00±0.31)$\times 10^{-3}$ | (0.8±0.12)$\times 10^{-3}$ | (0.9±0.15)$\times 10^{-3}$ | (1.4±0.3)$\times 10^{-3}$ | (0.9±0.09)$\times 10^{-3}$ |
| **CSF ptau181/Aβ42** | 0.02±0.013 | 0.009±0.001 | 0.02±0.007 | 0.036±0.012 | 0.008±0.002 |
| **Centiloid** | 22.51±26.25 | 0.93±8.30 | 25.81±21.32** | 46.84±20.55** | -6.23±6.76 |
| **Braak I/II SUVR** | 1.15±0.19 | 1.07±0.14 | 1.12±0.12 | 1.22±0.25 | 1.10±0.08 |
| **Braak III/IV SUVR** | 1.16±0.12 | 1.12±0.10 | 1.13±0.08 | 1.19±0.14 | 1.15±0.02 |
| **Braak V/VI SUVR** | 1.08±0.08 | 1.06±0.09 | 1.08±0.07 | 1.10±0.08 | 1.07±0.04 |
| **Global tau PET SUVR** | 1.11±0.09 | 1.08±0.09 | 1.10±0.07 | 1.13±0.10 | 1.10±0.03 |
| **CenTauR-z MetaROI** | 1.09±2.65 | 0.08±1.64 | 0.91±1.83 | 2.37±3.86* | 0.35±0.67 |
| **Hippocampal volume (mm<sup>3</sup>)** | 8860.14±1521.54 | 8566.00±1130.95 | 8670.81±831.13 | 8380.19±1219.52 | 9245.44±556.99 |
| **Z-Scored PACC, Mean (SD)** | 0.05±0.62 | 0.16±0.59 | 0.11±0.58 | -0.19±0.69 | 0.22±0.59 |

Based on the positivity threshold for each Braak stage, nine cases (9.09%) were positive in the Braak I/II region. Following a hierarchical pattern, five of them (5.05%) were also positive for the Braak III/IV ROI and one for the Braak V/VI region (Table_S2). However, two scans did not follow the hierarchical pattern and were positive only in the Braak III/IV ROI (Figure_S4). Both participants were in their 70s, APOE 4 carriers, female, with high level of Centiloid (89 and 78), and 8 years of education. Visual inspection of both scans confirmed an elevated level of tau PET signals in the Braak III/IV region, reassuring correct staging (Figure_S5).

In addition to SUVR, CenTauR-z in meta-ROI was calculated for all tau PET scans. Figure 1 illustrates the relationship between Centiloid values and both Braak I/II SUVR and CenTauR-z, stratified by CSF-based AT status. CenTauR-z and Braak I/II SUVR have a correlation of r=0.82 [95%CI: 0.60 to 0.92, p<0.001]. The correlation between Centiloid and Braak I/II SUVR was statistically significant only in the A+T+ group [r=0.53, 95% CI: 0.10–0.83, p=0.01]. Using a linear regression model, CenTauR-z = 2 was equivalent to 1.19 Braak I/II SUVR (Braak I/II = 1.09 + 0.05 × CenTauR-z), which is lower than the estimated positivity threshold and resulted in higher number of tau PET scans with CenTauR-z above 2.

**Figure 1.**
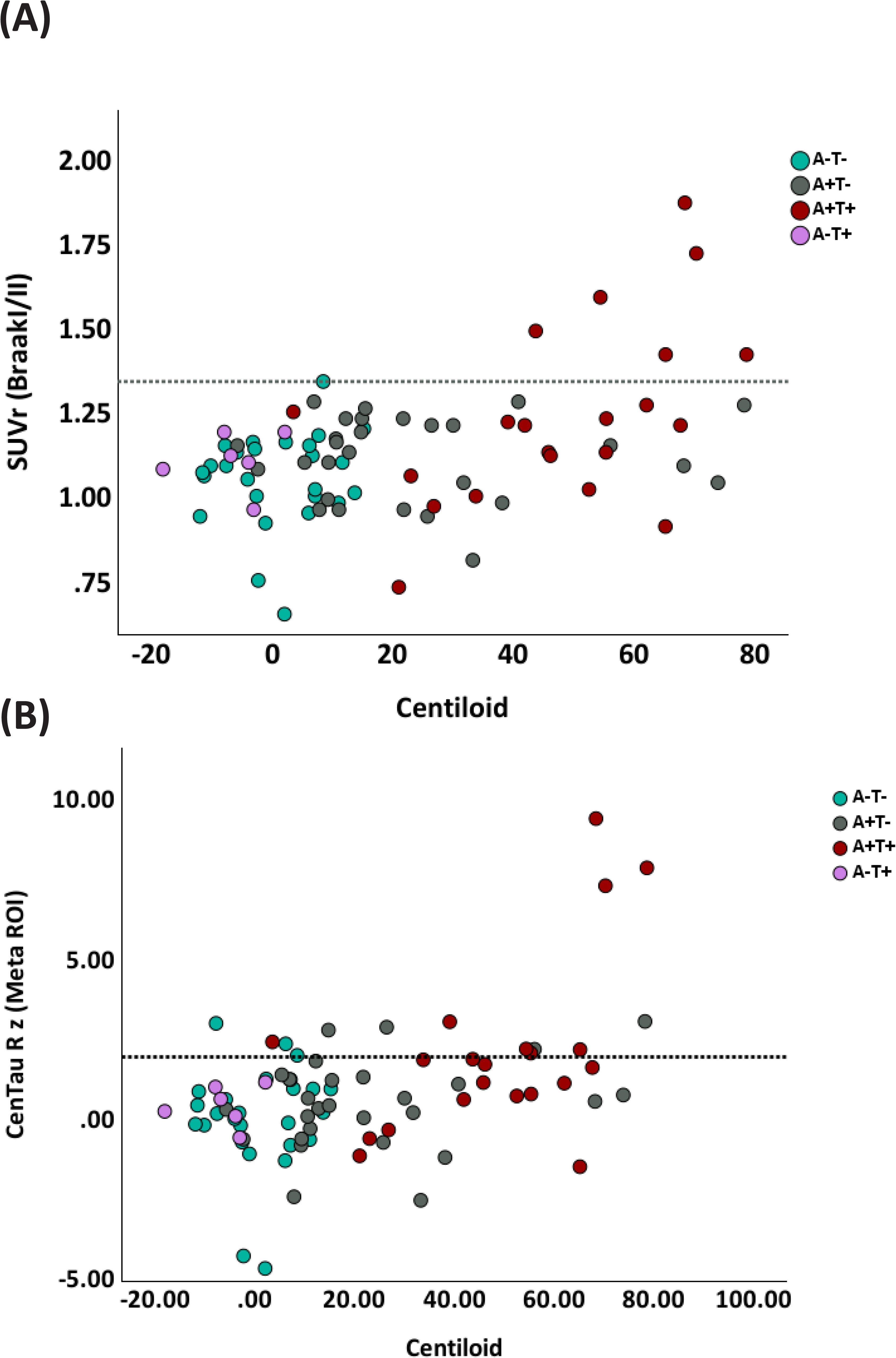
Scatter plot of Centiloid values (x-axis) and (A) Braak I/II tau PET SUVR (y-axis), (B) CenTauR-z (meta-ROI) with data points color-coded by PET-defined groups: A-T-(cyan), A+T-(gray), A+T+ (red), and A-T+ (purple). The dotted horizontal line represents the predefined SUVR threshold for tau positivity and z=2 for CenTauR-z, respectively. Higher Centiloid values are associated with increased tau PET SUVR, particularly in the A+T+ group (red), indicating a relationship between amyloid burden and early tau deposition in the Braak I/II region.

Figure 2 shows the partial correlations between Centiloid and tau PET Braak I/II with CSF- and plasma-based AD biomarkers, adjusting for age, sex, APOE ε4 status, and the time interval between PET imaging and fluid biomarkers collection. For the correlations between tau PET SUVR in Braak ROIs, the time interval was statistically significant, likely due to the longer time interval between measurements. In contrast, for Centiloid, the time interval was not statistically significant, as the plasma biomarkers were measured more concurrent to amyloid PET. CSF ptau217 showed a strong correlation with Centiloid [r=0.71, 95%CI: 0.61–0.77, p<0.001] and a moderate correlation with Braak I/II tau PET SUVR [r=0.58, 95%CI: 0.26–0.79, p<0.001]. Among other CSF biomarkers, CSF ptau181/Aβ42 also showed high correlations with both amyloid [r=0.72, 95%CI: 0.60–0.79, p<0.001] and Braak I/II tau PET [r=0.48, 95%CI: 0.22–0.69, p<0.001].

**Figure 2.**
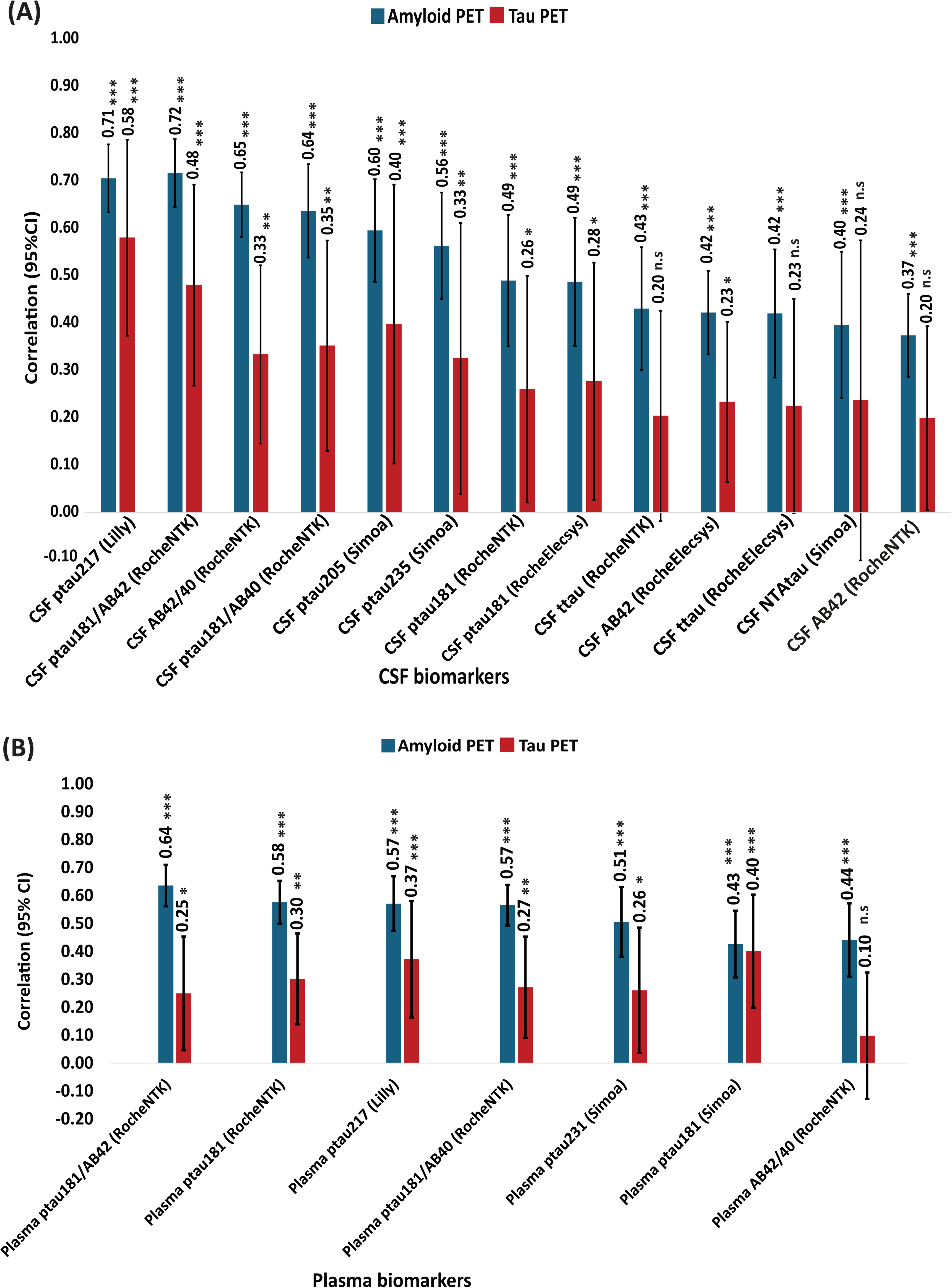
Partial correlations (with 95% confidence intervals and p-values) between amyloid burden (Centiloid values) and tau PET SUVR in Braak I/II regions and (A) CSF biomarkers, (B) plasma biomarkers. All analyses were adjusted for age, sex, APOE ε4 status, and the time interval between PET imaging and fluid biomarker collection.

Among the plasma biomarkers, ptau181/Aβ42 had the highest correlation with amyloid PET [r=0.64, 95%CI: 0.49–0.65, p<0.001], while ptau181 and ptau217 had the strongest correlations with Braak I/II SUVR [r=0.40, 95%CI: 0.12–0.60, p<0.001, and r=0.37, 95%CI: 0.09–0.58, p<0.001 respectively]. Additionally, plasma ptau181 [r=0.34, 95%CI: 0.07-0.60, p<0.001], ptau217 [r=0.29, 95%CI: 0.04-0.55, p<0.001] and ptau231 [r=0.27, 95%CI: 0.04-0.53, p<0.001] each showed moderate correlation with Braak III/IV tau PET SUVR. Among the CSF biomarkers, ptau217 showed the strongest correlation with Braak III/IV SUVR [r=0.46, 95%CI: 0.08-0.72, p<0.001], followed by ptau181/Aβ42 [r=0.38, 95%CI: 0.09-0.62, p<0.001] (Table_S3). None of the fluid biomarkers showed significant correlations with either Braak V/VI or global tau PET SUVR. Similarly, no statistically significant correlations were found between Braak I/II SUVR and cognitive measures.

Table_S4 and Table 2 present the performance of fluid biomarkers in predicting Braak I/II tau PET positivity, unadjusted and adjusted for age, sex, and APOE 4 carriership, respectively. Comparison of the AUCs between unadjusted and adjusted models indicates that biomarker performance (AUC and 95% CI) improves with adjustment; however, the overall pattern remains consistent for most biomarkers. Notably, the plasma Aβ42/40 ratio and CSF pTau235 do not demonstrate statistically significant performance in the unadjusted models. Table 2 summarizes the performance of fluid biomarkers and amyloid PET for predicting Braak I/II tau PET positivity, along with their respective positivity thresholds. Additionally, each biomarker’s PPV and NPV are reported. Models incorporating age, sex, and APOE 4 alone had an AUC of 0.78 [95%CI: 0.61–0.95, p < 0.005], serving as the baseline reference for biomarker evaluation. Adding fluid biomarkers/Centiloid to the model improved the performance and resulted in higher AUC for the biomarkers. In brief, among plasma biomarkers, ptau181 demonstrated the highest [AUC=0.86, 95%CI: 0.72– 1.00, p<0.0001]. Other plasma biomarkers, including ptau217 [AUC=0.84, 95%CI: 0.68– 0.99, p<0.001], and ptau181/Aβ42 [AUC=0.84, 95% CI: 0.65–0.99, p<0.001], also showed strong predictive performance. Among CSF biomarkers, ptau181/Aβ42 and ptau217 exhibited the highest AUC of 0.94 [95%CI: 0.85–1.00, p<0.0001] and 0.93 [0.95%CI: 0.85-1.00, p<0.0001], respectively.

**Table 2.**
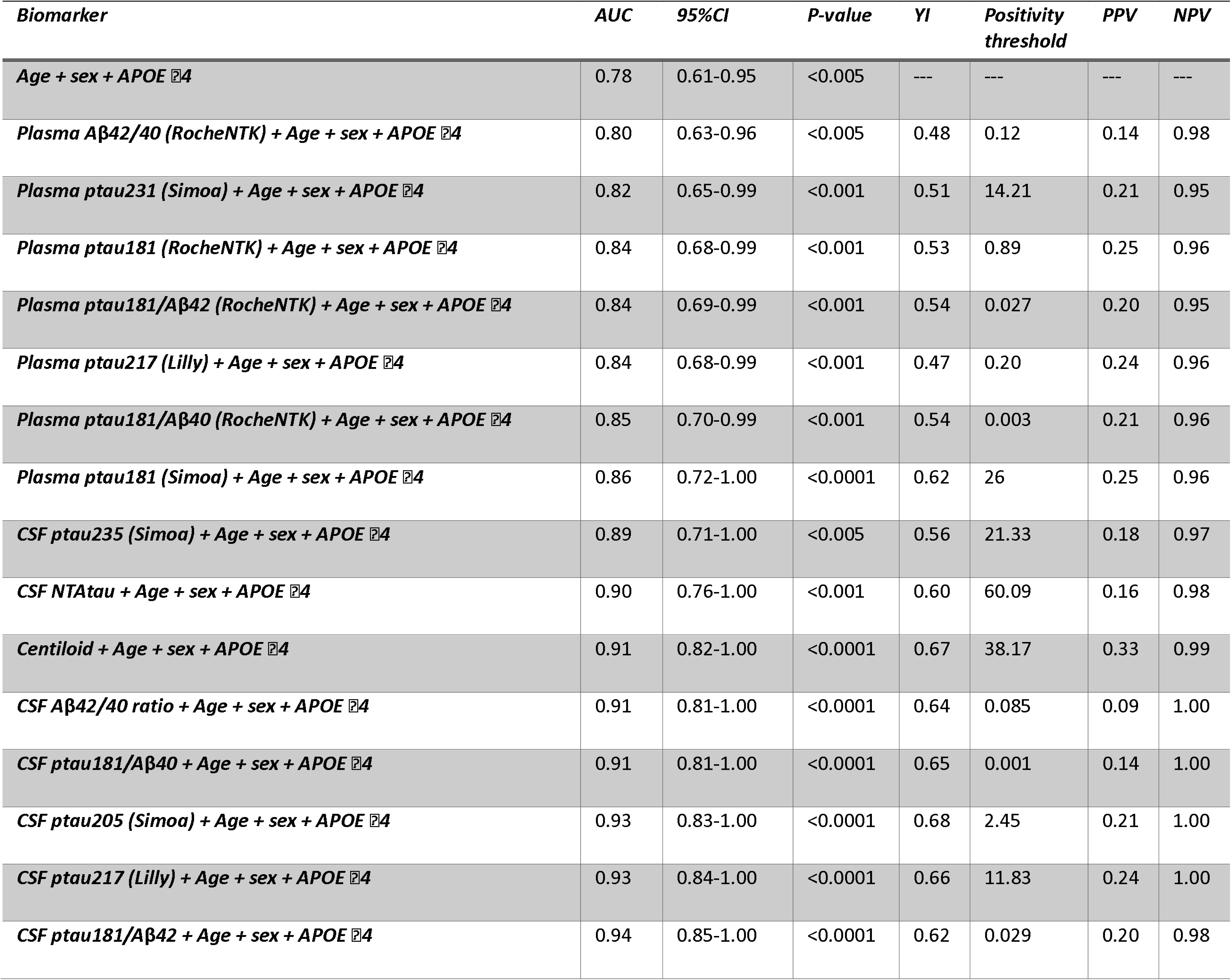
Area under curve (AUC; 95%CI), corresponding p-value, Youden Index (YI), positivity threshold, Positive Predictive Value (PPV), and Negative Predictive Value (NPV) of core AD fluid biomarkers and Centiloid for predicting Braak I/II positivity, extracted from ROC analysis.

PPV values for all biomarkers ranged from low to moderate, with Centiloid demonstrating the highest PPV (0.33), indicating slightly better predictive value for detecting Braak I/II positivity compared to fluid biomarkers, such as CSF and plasma ptau217 (PPV=0.24). Using our tau-PET staging, 90.9% of participants were classified as negative, and applying the 0.2 cut-off for plasma ptau217 similarly identified 96% as negative. This high proportion of tau-PET negative individuals likely accounts for the strong negative predictive values (NPVs) observed across all biomarkers, each exceeding 0.94.

To establish PET-based staging, we applied positivity thresholds derived from ROC analyses (Table 2). Amyloid PET positivity (A status) was defined using a Centiloid cut-off of 38.17, while tau PET positivity (T status) was determined using a Braak I/II SUVR threshold of 1.36. Participants with Centiloid values between 12 and 38.17 were classified as being in the amyloid PET gray zone (Gz). Based on these PET-derived thresholds, participants were grouped as follows:

PET (A-T-): Centiloid<12, and Braak I/II<1.35

PET (A(Gz)T-): 12≤Centiloid<38.17, and Braak I/II<1.35

PET (A+T-): Centiloid≥38.17, and Braak I/II<1.35

PET(A+T+): Centiloid≥38.17, and Braak I/II≥1.35

It is worth noting that, under the PET-based staging criteria, no participants were classified as PET (A–T+) or PET (A(Gz)T+). Demographic characteristics of the participants in each PET-based AT staging group are shown in Table_S5.

Figure 3 shows the comparison between fluid biomarkers, Centiloid, and cognition as a function of the PET-derived staging. Plasma biomarker levels demonstrated a stepwise increase across PET-defined groups, with the highest values observed in the A+T+ group. Significant differences between PET-defined stages were observed for both plasma ptau181 assays, and for plasma ptau217, demonstrating the pronounced stage-dependent elevations. Notably, plasma ptau181/Aβ42 also showed significant elevations in A+T+ individuals, reflecting their strong association with amyloid and tau pathology. CSF ptau217 and ptau181/Aβ42 presented the highest dynamic range and differentiation across different PET-based stages, followed by ptau235 and ptau205. Although cognitive scores showed a stepwise decline with progression to more advanced stages, the between-group differences did not reach statistical significance after applying Bonferroni correction for multiple comparisons (Figure 3-D). When comparing the fold changes across different plasma biomarkers, ptau181, ptau181/Aβ42, ptau217, ptau231, showed approximately 2-fold increase in the PET(A+T+) group relative to the reference (A-T-) group (Figure 4-A). Most CSF biomarkers showed increased fold changes across PET stages, with ptau217 and ptau181/Aβ42 having the highest increases about 4.5- and 5.5-fold in the PET(A+T+) group. NTAtau showed a ∼3-fold rise in the PET(A+T+) group compared to PET(A-T-) (Figure 4-B and 4-C). The statistical report for between group comparison for both z-scored biomarkers and ratio can be found in the supplementary materials, Table_S6 to Table_S10.

**Figure 3.**
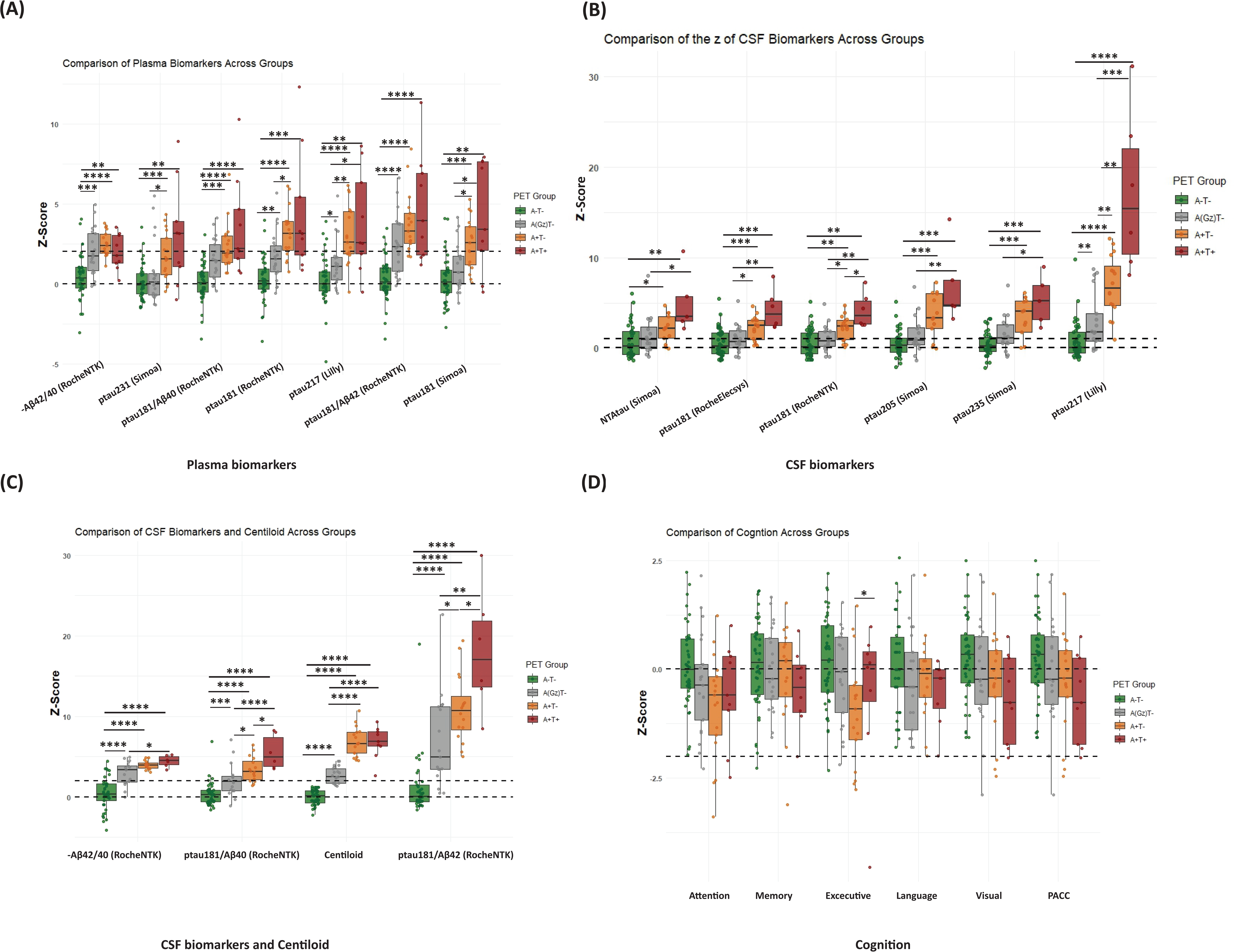
Group-wise comparisons of plasma, CSF, imaging biomarkers, and cognitive composites across PET-defined groups. (A) Plasma biomarker z-scores across PET(A-T-), PET(AGZT-), PET(A+T-), and PET(A+T+). (B) CSF ptau biomarker z-scores across the same groups. (C) Additional CSF and AD imaging biomarker z-scores across PET-defined groups. (D) Z-scored cognitive composite measures across the PET-defined groups. Statistical comparisons between groups are indicated, with significant differences denoted by p-values. ** Note that the z-scores of the Aβ42/40 ratios were multiplied by −1 for clearer presentation.

**Figure 4.**
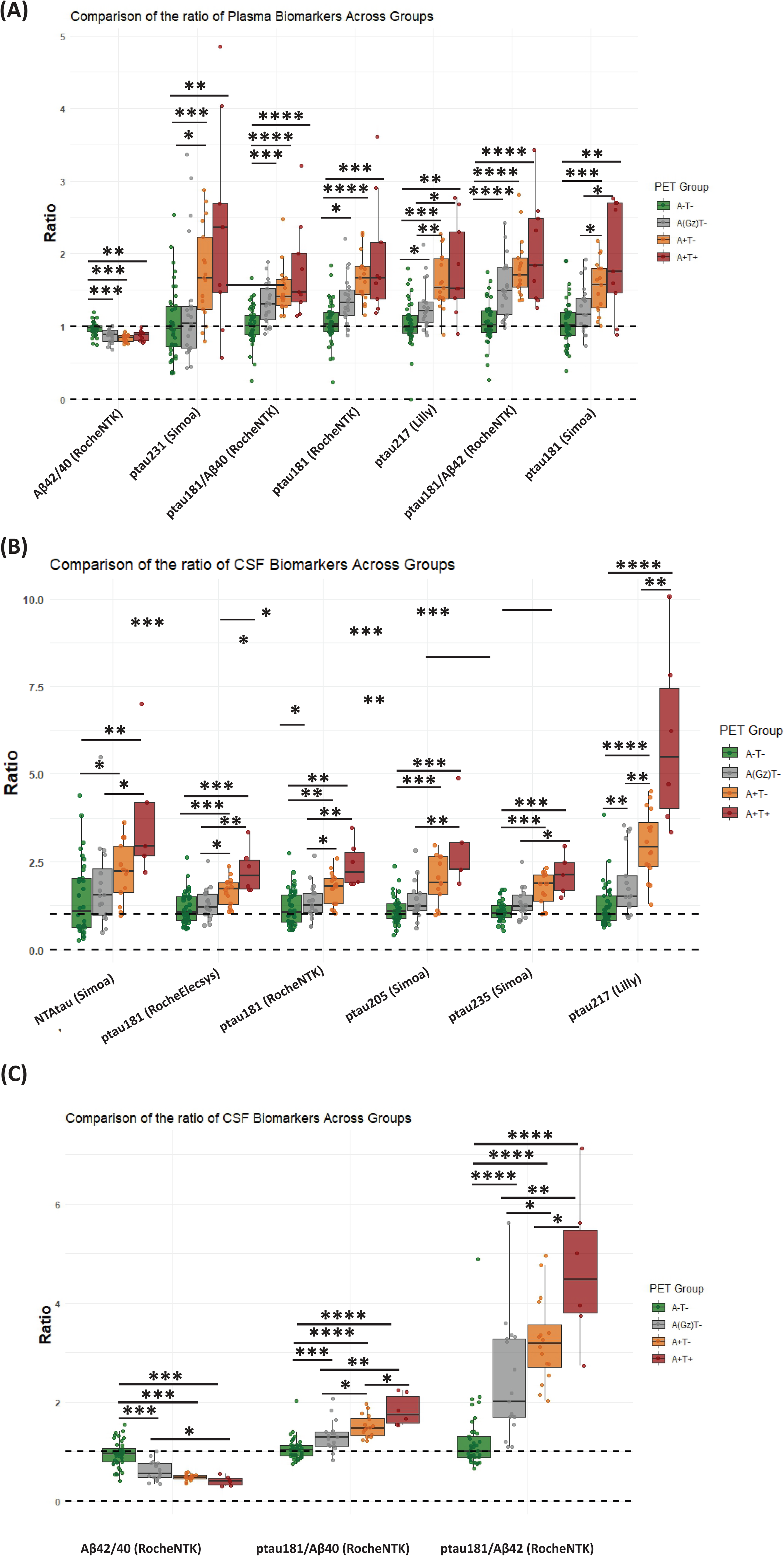
Group-wise comparisons of biomarker ratios relative to the PET-based reference group. The figure illustrates the fold changes in plasma and CSF biomarkers across PET-defined stages, highlighting significant differences between-groups.

## 4 DISCUSSION

In this study, we used tau PET to investigate NFT pathology in the earliest preclinical stages of the AD continuum and the associations with fluid-based AD biomarkers in plasma (Aβ42/40, ptau181, ptau181/Aβ42, ptau181/Aβ40, ptau217, and ptau231), and CSF (Aβ42/40, ptau181, ptau181/Aβ42, ptau181/Aβ40, ptau205, ptau235, and NTAtau). To comprehensively characterize these associations, we also examined relationships with amyloid pathology using Centiloid and cognitive performance.

Our findings reveal that tau PET signal is elevated during preclinical AD, with tau positivity in Braak I/II emerging in less than half of the individuals with Centiloid above 38.18. This suggests that neurofibrillary tangle (NFT) pathology can emerge earlier than previously recognized in the AD continuum, as prior reports have typically described tau positivity at Centiloid values exceeding 50 CL^44,45^. Moreover, staging of the ^18^F-RO-948 PET scans in our sample aligned with the Braak hierarchical staging model for most participants with tau positivity. Previous work has shown heterogeneity beyond Braak staging in the tau PET spread^13^. However, in our work, the ‘limbic’ subtype, which mostly corresponds with the pattern of tau spread in the Braak model, was associated with APOE ε4 carriership. The high prevalence of APOE ε4 carriers in our sample may explain the strong conformance to the Braak model observed in our cohort. However, observing 2 participants with the MTL-sparing tau pattern in our cohort reveals that distinct tau propagation patterns can already be detected even in the early stages. In addition to the common Braak staging, we used the CenTauR-z meta-ROI to detect tau PET positivity. Our results showed that the CenTauR-z method assigned a Z-score ≥ 2 to some tau PET scans from participants classified as A-T-and A+T-, despite their Centiloid values being low to moderate (CL < 30). This discrepancy between staging methods likely stems from methodological differences, including the use of distinct regions of interest (ROIs). Notably, our findings are consistent with those reported in the CenTauR-z study (PMID: 37424964), which demonstrated that CenTauR-z offers a robust quantitative framework for distinguishing between low and high levels of tau pathology^32^. Based on our results, for individuals at the early stage of tau accumulation, subject-specific Braak ROI quantifications for tau PET appears more sensitive in capturing subtle changes in tau deposition. Moreover, our tau PET quantification pipeline may further enhance early detection by accounting for meningeal uptake and minimizing off-target binding signal spillover into cortical target ROIs.

Plasma ptau biomarkers demonstrated a moderate association with Braak I/II tau PET SUVR, with ptau181 and ptau217 showing the strongest correlations, even after adjusting for age, sex, and APOE ε4 carriership. For amyloid PET Centiloid measures, plasma ptau181/Aβ42 exhibited the strongest association, followed by ptau181, ptau217, and ptau231, with the latter three showing comparable performance (r ∼ 0.58). As expected, CSF biomarkers had stronger correlations with both amyloid and tau PET signals, with CSF ptau217 showing the highest associations (r = 0.71 for amyloid PET and r = 0.58 for tau PET), followed by ptau181/Aβ42 (r=0.72 and r=0.48, respectively). Additionally, both plasma and CSF ptau217 showed moderate associations with Braak III/IV tau PET signal.

For fluid biomarkers and amyloid PET Centiloid, our ROC analysis showed good to excellent AUCs when entering age, sex and APOE ε4 status in the model. Despite high NPV of all biomarkers (>0.94), their PPV remained low, with Centiloid achieving the highest PPV (0.33), followed by plasma and CSF ptau biomarkers (∼0.24–0.25). This is probably due to the low prevalence of tau PET positivity in our cohort, where a high proportion of tau-negative cases inflates the NPV and AUC. Applied to screening tau PET-positive cognitively unimpaired individuals for a clinical trial, this means that only 25% of the participants predicted to be positive by the best-performing plasma biomarkers studied here are expected to actually be tau PET positive. On the other hand, when combined with demographic and genetic information, plasma biomarkers are very accurate (>95%) at ruling out the tau PET positivity, which may be useful for diagnostic purposes, for instance in individuals with subjective cognitive complaints.

We classified participants into four distinct groups based on their amyloid and tau PET status to evaluate the ability of fluid biomarkers to track the progression of fibrillar core-AD pathology. Importantly, all participants in our cohort adhered to this classification, with none being tau PET-positive while amyloid-negative or in the amyloid gray zone, ensuring alignment with the AD continuum. It should be noted that our positivity threshold was defined as the mean plus two standard deviations (mean + 2SD) of regional Braak ROI uptake in a reference group of cognitively unimpaired, middle-aged individuals who were A-T-based on CSF biomarkers. The rationale for selecting the reference group based on CSF AT status lies in the expectation that CSF biomarkers become abnormal earlier than PET measures, making this a reasonable and sensitive approach to defining abnormality^46^. The consistency of this method is further supported by the resulting staging: no individuals classified as CSF A-T-were tau PET-positive, and all tau PET-positive individuals had Centiloid values above 38. Importantly, the design of our staging method did not exclude amyloid PET negative, tau PET positive cases by definition, as the thresholds for abnormality were determined independently for CSF-based amyloid and tau biomarkers. Our findings highlight that CSF ptau217 and ptau181/Aβ42 exhibited the greatest dynamic range and the largest fold increases across PET-based AD stages, with a clear stepwise rise as pathology progressed. Among plasma biomarkers, ptau181, ptau181/Aβ42, and ptau217 also showed stepwise increases across stages, though with lower fold changes compared to their CSF counterparts.

Furthermore, our analysis of biomarker fold changes across PET-based stages demonstrated that CSF ptau217 and ptau181/Aβ42 had the most pronounced increases, particularly in the PET(A+T+) stage, reinforcing their strong association with tau pathology and amyloid plaques. Although plasma ptau217 showed weaker performance than its CSF counterpart, it still exhibited a ∼1.5-fold increase in the more advanced stages of pathology compared to PET(A-T-), confirming its alteration due to the core-AD pathology.

These results were obtained in a sample with a relatively balanced distribution of participants in each CSF-based AT stage, as determined by study design. It is essential to highlight that participants in this study are in the early stage of AD, all classified as CU individuals. Their mean age (65.15±5.00), as well as their amyloid load measured by PET (Centiloid=22.52±26.25), is considerably lower than in other studies found in the literature that include CU individuals. These results support that the sample studied is at relatively early stages of the preclinical AD continuum and can also explain the lack of associations observed between tau PET and cognitive performance. This sample, being at the earliest preclinical AD continuum, can be considered a strength of the study. In addition, participants have been thoroughly characterized with several fluid and imaging biomarkers, showcasing the interrelationships of these biomarkers in the earliest pathological AD continuum.

On the other hand, this study is not free of limitations. This sample is highly selected and enriched for AD risk factors, particularly amyloid, as reflected in the high percentage of APOE ε4 carriers. While this may influence tau prevalence estimates and the detection of certain tau subtypes compared to unselected populations, it is unlikely to significantly affect the associations between tau PET and other AD biomarkers, which should remain broadly generalizable. Moreover, the cross-sectional nature of the analyses does not allow us to study several interesting questions, such as the progression of tau spread over time or the capacity of fluid biomarkers to predict tau accumulation in the future. In this regard, the longitudinal collection of ^18^F-RO-948 in this sample is underway and will allow us to address these questions in the near future.

To summarize, ^18^F-RO-948 PET was capable of detecting positive cases in participants of the ALFA+ cohort who are cognitively unimpaired and at the earliest preclinical AD stages. Most of the positive cases conformed to the Braak hierarchical model. Despite the association between the tau PET SUVRs and fluid biomarkers, the fluid biomarkers studied here, and more specifically plasma biomarkers, show limited capacity to detect tau PET positivity (∼25%) and, therefore, have a restricted capacity to screen tau PET positive participants for prevention trials. On the other hand, plasma biomarkers display a very high capacity to rule out tau PET positivity, which may bring diagnostic value.

## Supporting information

Supplementary Matterial

## Data Availability

All requests for raw and analyzed data and materials will be promptly reviewed by the corresponding authors and the Barcelonaβeta Brain Research Center to verify whether the request is subject to any intellectual property or confidentiality obligations. Bulk Anonymized data can be shared by request from any qualified investigator for the sole purpose of replicating procedures and results presented in the article, provided that data transfer is in agreement with EU legislation.

## ACKNOWLEDGEMENTS

This publication is part of the BBRC’s ALFA study. The authors would like to express their most sincere gratitude to the project participants, without whom this research would not have been possible. Collaborators of the ALFA study are: Müge Akinci, Federica Anastasi, Annabella Beteta, Raffaele Cacciaglia, Lidia Canals, Alba Cañas, Carme Deulofeu, Maria Emilio, Irene Cumplido-Mayoral, Marta del Campo, Carme Deulofeu, Ruth Dominguez, Maria Emilio, Karine Fauria, Sherezade Fuentes, Marina García, Laura Hernández, Gema Huesa, Jordi Huguet, Laura Iglesias, Esther Jiménez, David López-Martos, Paula Marne, Tania Menchón, Paula Ortiz-Romero, Eleni Palpatzis, Wiesje Pelkmans, Albina Polo, Sandra Pradas, Mahnaz Shekari, Lluís Solsona, Anna Soteras, Núria Tort-Colet, and Marc Vilanova.

The authors thank F. Hoffmann-La Roche Ltd for sponsoring the tau PET study, Roche Diagnostics International Ltd for providing the kits to measure CSF biomarkers, Eli Lilly and Company for providing the measurements of the in-house assay for CSF and plasma ptau217, and GE Healthcare for providing the doses of 18F-flutemetamol PET. The NeuroToolKit is a panel of exploratory prototype assays designed to robustly evaluate biomarkers associated with key pathologic events characteristic of AD and other neurological disorders, used for research purposes only and not approved for clinical use (Roche Diagnostics International Ltd, Rotkreuz, Switzerland). Elecsys β-amyloid (1–42) CSF, Elecsys Phospho-Tau (181P) CSF, and Elecsys Total-Tau CSF assays are approved for clinical use. ELECSYS is a trademark of Roche. All other product names and trademarks are the property of their respective owners.

## CONFLICT OF INTEREST STATEMENT

Theresa A Day is a full-time employee and stockholder of Eli Lilly and Company, Indianapolis, IN, USA. Jeffrey L Dage (JLD) is an inventor on patents or patent applications assigned to Eli Lilly and Company relating to the assays, methods, reagents and/or compositions of matter for P-tau assays and Aβ targeting therapeutics. JLD has/is served/serving as a consultant or on advisory boards for Eisai, Abbvie, Genotix Biotechnologies Inc, Gates Ventures, Gate Neurosciences, Dolby Family Ventures, Karuna Therapeutics, Alzheimer’s Disease Drug Discovery Foundation, ALZpath Inc., Cognito Therapeutics, Inc., Eli Lilly and Company, Prevail Therapeutics, Neurogen Biomarking, Spear Bio, Rush University, University of Kentucky, Tymora Analytical Operations, and Quanterix. JLD has received research support from ADx Neurosciences, Fujirebio, Roche Diagnostics, and Eli Lilly and Company in the past two years. JLD has received speaker fees from Eli Lilly and Company and LabCorp. JLD is a founder and advisor for Monument Biosciences and Dage Scientific LLC. JLD has stock or stock options in Eli Lilly and Company, Genotix Biotechnologies, AlzPath Inc., Neurogen Biomarking, and Monument Biosciences.

Thomas Karikari (TKK) has consulted for Quanterix Corporation, SpearBio Inc., Neurogen Biomarking LLC., and Alzheon, and has served on advisory boards for Siemens Healthineers, Neurogen Biomarking LLC., and Alzheon (which may come with minority stock equity interest/stock options), outside the submitted work. He has received in-kind research support from Janssen Research Laboratories, SpearBio Inc., and Alamar Biosciences, as well as meeting travel support from the Alzheimer’s Association and Neurogen Biomarking LLC., outside the submitted work. TKK has received royalties from Bioventix for the transfer of specific tau antibodies and assays to third-party organizations. He has received honoraria for speaker/grant review engagements from the NIH, UPENN, UW Madison, the Cherry Blossom symposium, the HABS-HD/ADNI4 Health Enhancement Scientific Program, Advent Health Translational Research Institute, Brain Health conference, Barcelona Pittsburgh conference, the International Neuropsychological Society, the Icahn School of Medicine at Mount Sinai and the Quebec Center for Drug Discovery, Canada, all outside of the submitted work. TKK serves/has served as a guest editor for npj Dementia, as an invited member of the World Health Organization committee to develop preferred product characteristics for blood-based biomarker diagnostics for Alzheimer’s disease, as an executive committee member for the Human Amyloid Imaging (HAI) conference, as an elected member of the NACC ADRCs Steering Committee, as co-director of the NACC ADRCs Biofluid Biomarker Working Group, and as a member of the Alzheimer’s Association committees to develop Appropriate Use Criteria for clinical use of blood-based biomarkers, and treatment related amyloid clearance. TKK is an inventor on several patents and provisional patents regarding biofluid biomarker methods, targets, and reagents/compositions, that may generate income for the institution and/or self should they be licensed and/or transferred to another organization. These include WO2020193500A1: Use of a ps396 assay to diagnose tauopathies; 63/679,361: Methods to Evaluate Early-Stage Pre-Tangle TAU Aggregates and Treatment of Alzheimer’s Disease Patients; 63/672,952: Method for the Quantification of Plasma Amyloid-Beta Biomarkers in Alzheimer’s Disease; 63/693,956: Anti-tau Protein Antigen Binding Reagents; and 2450702-2: Detection of oligomeric tau and soluble tau aggregates.

Matteo Tonietto, Edilio Borroni, and Gregory Klein are full-time employees of F. Hoffmann - La RocheLtd., Basel, Switzerland. Margherita Carboni and Clara Quijano-Rubio are full-time employees of Roche Diagnostics International Ltd, Rotkreuz, Switzerland. Gwendlyn Kollmorgen is a full-time Roche Diagnostics GmbH employee. Eugeen Vanmechelen is a full-time employee of ADx NeuroSciences, Ghent, Belgium. José Luis Molinuevo is a full-time employee of Novartis, Basel, Switzerland. Henrik Zetterberg (H.Z.) is a Wallenberg Scholar and a Distinguished Professor at the Swedish Research Council supported by grants from the Swedish Research Council (#2023-00356, #2022-01018 and #2019-02397), the European Union’s Horizon Europe research and innovation programme under grant agreement No 101053962, Swedish State Support for Clinical Research (#ALFGBG-71320), the Alzheimer Drug Discovery Foundation (ADDF), USA (#201809-2016862), the AD Strategic Fund and the Alzheimer’s Association (#ADSF-21-831376-C, #ADSF-21-831381-C, #ADSF-21-831377-C, and #ADSF-24-1284328-C), the European Partnership on Metrology, co-financed from the European Union’s Horizon Europe Research and Innovation Programme and by the Participating States (NEuroBioStand, #22HLT07), the Bluefield Project, Cure Alzheimer’s Fund, the Olav Thon Foundation, the Erling-Persson Family Foundation, Familjen Rönströms Stiftelse, Stiftelsen för Gamla Tjänarinnor, Hjärnfonden, Sweden (#FO2022-0270), the European Union’s Horizon 2020 research and innovation programme under the Marie Skłodowska-Curie grant agreement No 860197 (MIRIADE), the European Union Joint Programme – Neurodegenerative Disease Research (JPND2021-00694), the National Institute for Health and Care Research University College London Hospitals Biomedical Research Centre, the UK Dementia Research Institute at UCL (UKDRI-1003), and an anonymous donor. H.Z. has served at scientific advisory boards and/or as a consultant for Abbvie, Acumen, Alector, Alzinova, ALZpath, Amylyx, Annexon, Apellis, Artery Therapeutics, AZTherapies, Cognito Therapeutics, CogRx, Denali, Eisai, Enigma, LabCorp, Merry Life, Nervgen, Novo Nordisk, Optoceutics, Passage Bio, Pinteon Therapeutics, Prothena, Quanterix, Red Abbey Labs, reMYND, Roche, Samumed, Siemens Healthineers, Triplet Therapeutics, and Wave, has given lectures sponsored by Alzecure, BioArctic, Biogen, Cellectricon, Fujirebio, Lilly, Novo Nordisk, Roche, and WebMD, and is a co-founder of Brain Biomarker Solutions in Gothenburg AB (BBS), which is a part of the GU Ventures Incubator Program (outside submitted work). Kaj Blennow is supported by the Swedish Research Council (#2017-00915 and #2022-00732), the Swedish Alzheimer Foundation (#AF-930351, #AF-939721 and #AF-968270), Hjärnfonden, Sweden (#FO2017-0243 and #ALZ2022-0006), the Swedish state under the agreement between the Swedish government and the County Councils, the ALF-agreement (#ALFGBG-715986 and #ALFGBG-965240), the Alzheimer’s Association 2021 Zenith Award (ZEN-21-848495), and the Alzheimer’s Association 2022-2025 Grant (SG-23-1038904 QC). Marc Suárez-Calvet receives funding from the European Research Council (ERC) under the European Union’s Horizon 2020 research and innovation programme (Grant agreement No. 948677), Project “PI19/00155”, funded by Instituto de Salud Carlos III (ISCIII) and co-funded by the European Union, and from a fellowship from “la Caixa” Foundation (ID 100010434) and from the European Union’s Horizon 2020 research and innovation programme under the Marie Skłodowska-Curie grant agreement No 847648 (LCF/BQ/PR21/11840004). Thomas K. Karikari was supported by grants 1 R01 AG083874-01 and 1U24AG082930 from the National Institutes of Health (NIH), the Swedish Research Council (Vetenskåpradet; 2021-03244), the Alzheimer’s Association (AARF-21-850325), the Swedish Alzheimer Foundation (Alzheimerfonden), the Aina (Ann) Wallströms and Mary-Ann Sjöbloms Stiftelsen, and the Emil och Wera Cornells stiftelsen. Juan Domingo Gispert (JDG) was supported by the Spanish Ministry of Science and Innovation (RYC 2013 13054). JDG has also received research support from the EU/EFPIA Innovative Medicines Initiative Joint Undertaking AMYPAD (grant agreement 115952), EIT Digital (grant 2021), and from Ministerio de Ciencia y Universidades (grant agreement RTI2018 102261). JDG is currently full-time employee at AstraZeneca. Gonzalo Sánchez-Benavides receives funding from the Ministerio de Ciencia e Innovación, Spanish Research Agency, PID2020-119556RA-I00; and the grant CP23/00039, funded by the Instituto de Salud Carlos III (ISCIII) and co-funded by the European Union/FSE+. Oriol Grau-Rivera receives funding from the Alzheimer’s Association Research Fellowship Program (2019-AARF-644568), from Instituto de Salud Carlos III (PI19/00117); the Spanish Ministry of Science, Innovation and Universities (Juan de la Cierva programme IJC2020-043417-I); and the grant IJC2020-043417-I, funded by MCIN/AEI/10.13039/501100011033 and the European Union NextGenerationEU/PRTR. Anna Brugulat-Serrat receives funding from the Alzheimer’s Association through the Alzheimer’s Association Clinician Scientist Fellowship (AACSF-23-1145154).

## SOURCE OF FUNDING

The ALFA+ study receives funding from “la Caixa” Foundation (project code LCF/PR/SC22/68000001), the Alzheimer’s Association, and an international anonymous charity foundation through the TriBEKa Imaging Platform project (TriBEKa 17 519007). Additional support has been received from the Universities and Research Secretariat, Ministry of Business and Knowledge of the Catalan government under grant no. 2021 SGR 00913.

This study has received financial support from F. Hoffmann-La Roche Ltd., with the project title “Tau imaging in the ALFA+ cohort study”.

## ROLE OF FUNDER/SPONSOR

The funders had no role in the design and conduct of the study, the collection, management, analysis, and interpretation of the data, the preparation, review, or approval of the manuscript, or the decision to submit the manuscript for publication.

## DISCLAIMER

This article reflects the views of the authors, and the Associations are liable for any use that may be made of the information contained herein.

## CONSENT STATEMENT

The ALFA study was conducted in accordance with the directives of the Spanish Law 14/2007, of July 3, on Biomedical Research (Ley 14/2007 de Investigación Biomédica). The ALFA study protocol was approved by the Independent Ethics Committee Parc de Salut Mar, Barcelona, and registered at Clinicaltrials.gov (Identifier: NCT01835717). All participants accepted the study procedures by signing the study’s informed consent form, which had also been approved by the same institutional review board.

## Notes

### Clinical Trial

NCT02485730

### Author Declarations

The Ethics Committee of Parc de Salut Mar (Barcelona, Spain) gave ethical approval for this work. The study was registered at Clinicaltrials.gov with the Identifier: NCT02485730.

