## Supplementary Matterial for "Associations of ^18^F-RO-948 Tau PET with Fluid AD Biomarkers, Centiloid, and Cognition in the Early AD Continuum"

^1^Barcelonaβeta Brain Research Center (BBRC), Pasqual Maragall Foundation. Barcelona, Spain.
^2^Hospital del Mar Research Institute (HMRI), Barcelona, Spain.
^3^Universitat Pompeu Fabra, Barcelona, Spain.
^4^Centro de Investigación Biomédica en Red de Fragilidad y Envejecimiento Saludable (CIBERFES), Instituto de Salud Carlos III, Madrid, Spain.
^5^Centro de Investigación Biomédica en Red de Bioingeniería, Biomateriales y Nanomedicina; (CIBERBBN), Instituto de Salud Carlos III, Madrid, Spain.
^6^Global Brain Health Institute, San Francisco, CA, USA.
^7^Nuclear Medicine Department, Hospital Clínic Barcelona - IDIBAPS, Spain.
^8^F. Hoffmann-La Roche Ltd, Basel, Switzerland.
^9^Department of Psychiatry and Neurochemistry, Institute of Neuroscience & Physiology, the Sahlgrenska Academy at the University of Gothenburg, Mölndal, Sweden^10^ Banner Alzheimer's Institute and University of Arizona, Phoenix, AZ, USA
^11^ Banner Sun Health Research Institute, Sun City, AZ 85351, USA
^12^Department of Psychiatry, University of Pittsburgh, PA, USA.
^13^Turku PET Centre, University of Turku, Turku, Finland.
^14^Lilly Research Laboratories, Eli Lilly and Company, Indianapolis, IN, USA.
^15^Stark Neurosciences Research Institute, Indiana University School of Medicine, Indianapolis, IN, USA.
^16^Roche Diagnostics GmbH, Penzberg, Germany.
^17^Roche Diagnostics International Ltd, Rotkreuz, Switzerland.
^18^ADx NeuroSciences, Technologiepark 94, Ghent, Belgium.
^19^Novartis Institutes for BioMedical Research, Translational Medicine, Neuroscience, Basel, Switzerland
^20^Department of Psychiatry and Neurochemistry, Institute of Neuroscience and Physiology, University of Gothenburg, Mölndal, Sweden.
^21^Clinical Neurochemistry Laboratory, Sahlgrenska University Hospital, Mölndal, Sweden.
^22^Department of Neurodegenerative Disease, UCL Institute of Neurology, Queen Square, London, United Kingdom.
^23^UK Dementia Research Institute at UCL, London, United Kingdom.
^24^Servei de Neurologia, Hospital del Mar, Barcelona, Spain.
^25^AstraZeneca

|  | *Total* | *A^-^T^-^* | *A^+^T^-^* | *A^+^T^+^* | *A^-^T^+^* |
| --- | --- | --- | --- | --- | --- |
| *N* | 212 | 86 | 51 | 26 | 9 |
| *Age, Mean (SD) [53-78 years]* | 65.67±4.85 | 64.05±4.58 | 64.16±4.75 | 67.38±4.98 | 65.39±2.31 |
| *Sex, N (%) [Female]* | 57 (57.57%) | 14 (51.85%) | 21 (63.63%) | 18 (64.28%) | 3 (33.33%) |
| *APOE-ε4, N (%) [Carriers]* | 63 (63.63%) | 12 (44.44%) | 26 (78.78%) | 19 (67.85%) | 4 (44.44%) |
| *CSF Aβ42/40, Mean (SD)* | 0.07±0.02 | 0.09±0.01 | 0.05±0.01 | 0.04±0.01 | 0.11±0.02 |
| *CSF ptau181, Mean (SD), pg/mL* | 18.49±8.35 | 14.50±4.06 | 16.53±3.95 | 31.98±8.55 | 30.11±8.29 |
| *CSF ptau181/Aβ40* | 0.0009±0.0003 | 0.0008±0.00014 | 0.0009±0.00015 | 0.0014±0.0003 | 0.001±0.0001 |
| *CSF ptau181/Aβ42* | 0.02±0.01 | 0.010±0.002 | 0.020±0.007 | 0.040±0.013 | 0.010±0.003 |
| *Centiloid* | 9.87±22.96 | -2.13±7.16 | 15.53±21.48 | 43.21±21.46 | -5.68±6.21 |

Table_S1. Demographic information, Centiloid, AD CSF-core biomarkers of participants stratified by CSF-based amyloid (A) and tau (T) status. All biomarkers in this table were measured within 3.00±4.08 months of the amyloid PET scan. Participants were categorized into AT stages using pre-established cut-offs^1^ based on CSF Aβ42/40<0.071 and p-tau181>24(pg/ml) measured with the NeuroToolKit, a panel of robust prototype assays and Elecsys® immunoassays (Roche Diagnostics International Ltd).

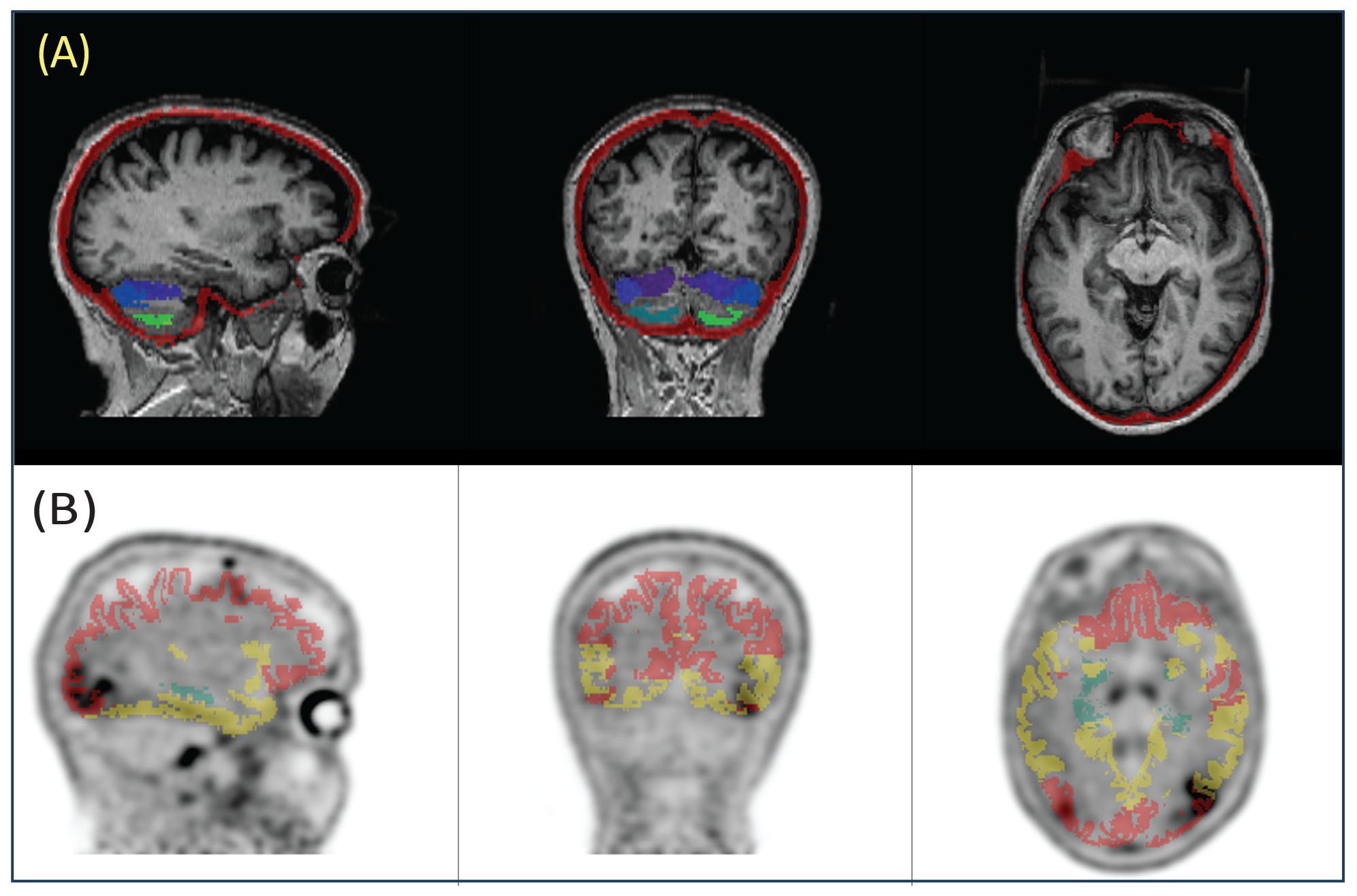

Figure_S1. (A) Fused T1-weighted MRI with meningeal mask and inferior cerebellum as reference region (B) Tau PET scan fused with the subject based cortical ROI, representing different Braak stages.

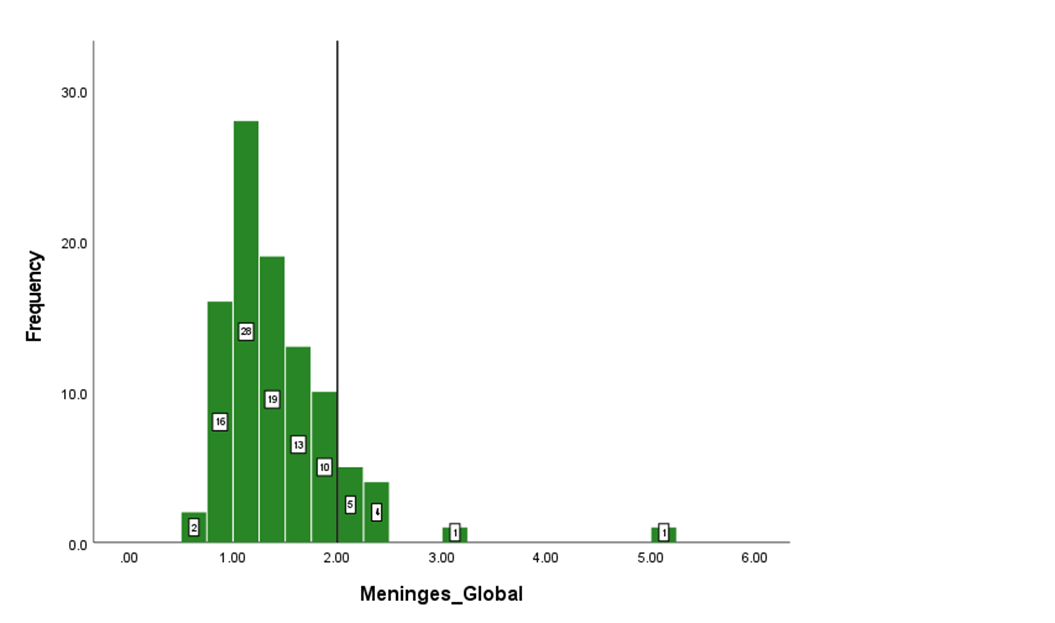

Figure_S2. Histogram of the global meningeal SUVr. It should be noted that 11 participants showed meningeal SUVr above 2.

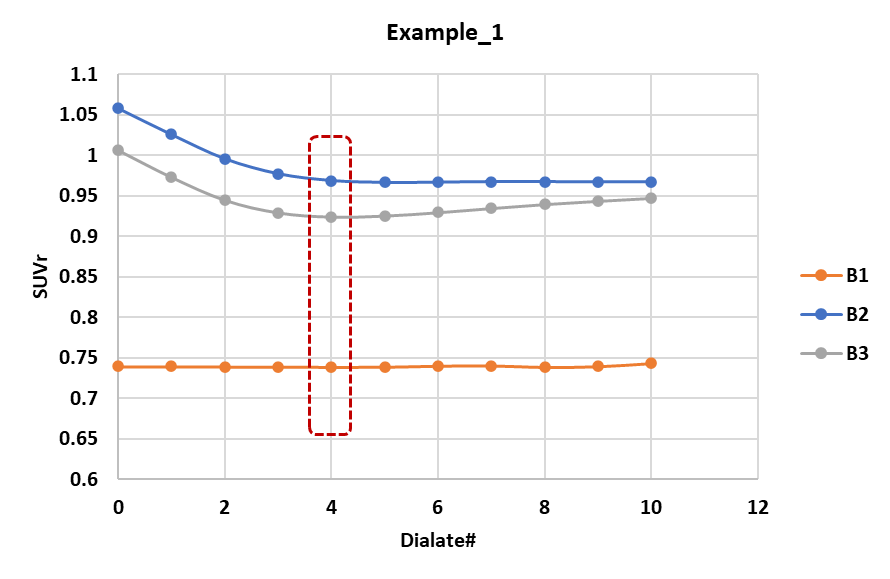

Figure_S3. An example of the procedure of choosing optimal SUVr for the 11 participants with high meningeal uptake. B1, B2, and B3 in the figure refer to Braak I/II, Braak III/IV, and Braak V/VI respectively.

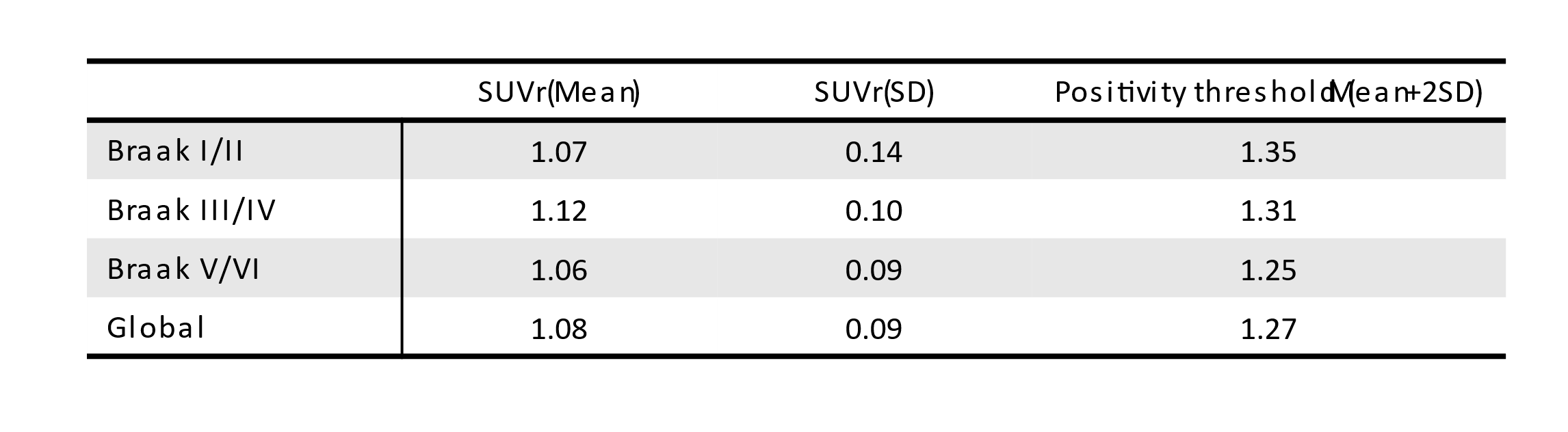

Table_S2. The positivity threshold for each Braak ROI calculated based on the mean plus two standard deviations extracted from CSF-based A-T-.

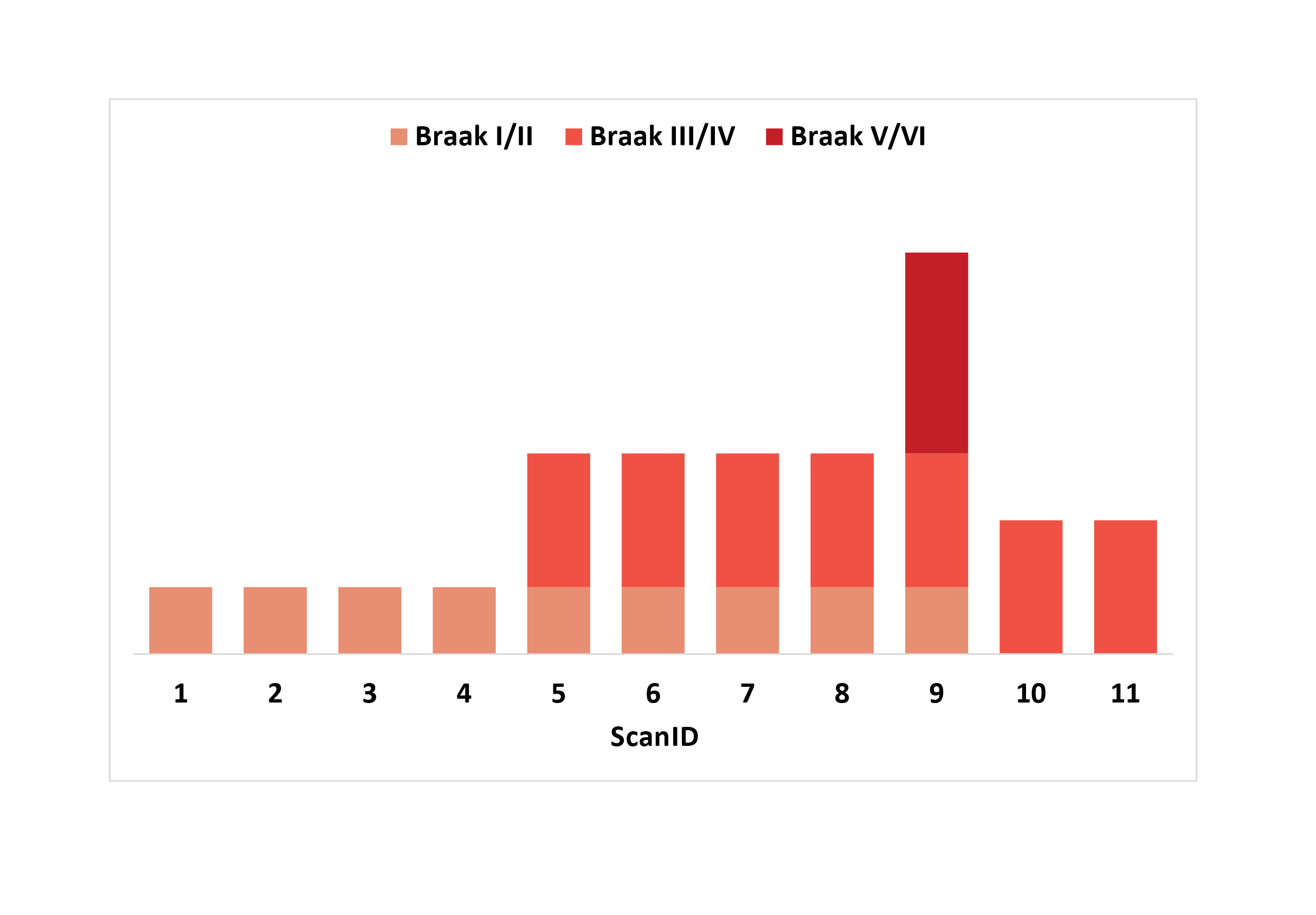
*Figure_S4. hierarchal presentation of tau PET positivity for each Braak stage.*

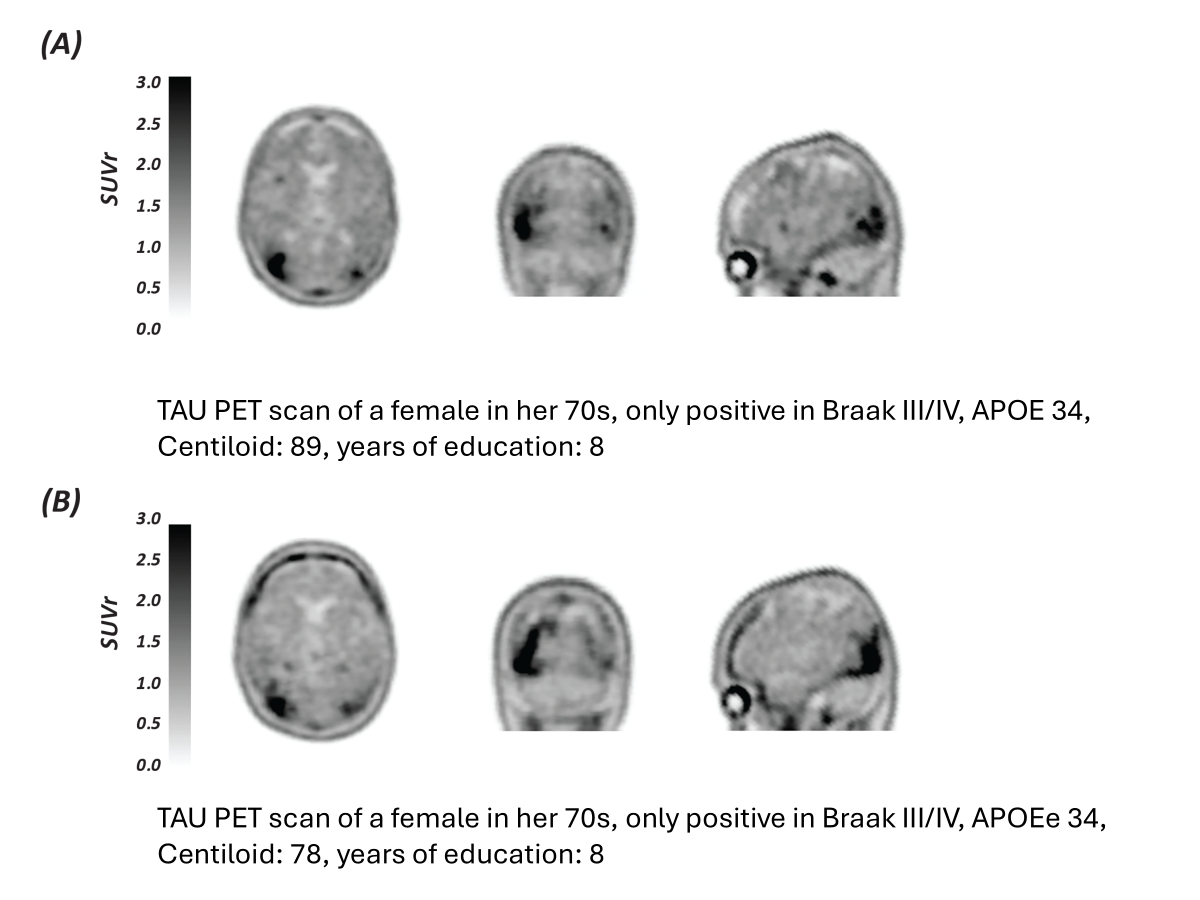

Figure_S5. Visual inspections of the tau PET scans deviating from Braak staging hierarchy, and their corresponding demographical information.

| Fluid biomarkers | Partial_Correlation | 95%CI | p_value |
| --- | --- | --- | --- |
| **Plasma ptau181 (Simoa)** | **0.34** | **0.07-0.60** | **0.001** |
| **Plasma ptau217 (Lilly)** | **0.29** | **0.04-0.55** | **0.005** |
| **Plasma ptau231 (Simoa)** | **0.27** | **0.04-0.54** | **0.009** |
| Plasma ptau181 (RocheNTK) | 0.18 | -0.01-0.37 | n.s |
| Plasma ptau181/Aβ40 (RocheNTK) | 0.17 | -0.03-0.39 | n.s |
| Plasma ptau181/Aβ42 (RocheNTK) | 0.17 | -0.06-0.41 | n.s |
| Plasma Aβ42/40 (RocheNTK) | -0.08 | -0.34-0.20 | n.s |
| **CSF ptau217 (Lilly)** | **0.46** | **0.08-0.73** | **<0.0001** |
| **CSF ptau181/Aβ42 (RocheNTK)** | **0.38** | **0.09-0.62** | **<0.0001** |
| CSF ptau205 (Simoa) | 0.32 | -0.03-0.66 | n.s |
| CSF ptau181/Aβ40 (RocheNTK) | 0.28 | 0.00-0.54 | n.s |
| **CSF Aβ42/40 (RocheNTK)** | **0.25** | **0.01-0.46** | **0.03** |
| CSF NTAtau (Simoa) | 0.24 | -0.07-0.57 | n.s |
| CSF ptau181 (RocheElecsys) | 0.24 | -0.04-0.49 | n.s |
| CSF ptau235 (Simoa) | 0.23 | -0.07-0.54 | n.s |
| CSF ptau181 (RocheNTK) | 0.22 | -0.05-0.47 | n.s |
| CSF ttau (RocheElecsys) | 0.21 | -0.05-0.43 | n.s |
| CSF ttau (RocheNTK) | 0.19 | -0.07-0.40 | n.s |
| CSF Aβ42 (RocheNTK) | -0.14 | -0.35-0.09 | n.s |
| CSF Aβ42 (RocheElecsys) | -0.16 | -0.35-0.06 | n.s |

Table_S3. Partial correlations and corresponding 95% confidence intervals between Braak III/IV SUVr and fluid biomarkers, adjusted for age, sex, APOE-ε4 carriership, and the time interval between PET imaging and fluid biomarker measurements.

| Biomarker | AUC | 95%CI | P-value |
| --- | --- | --- | --- |
| CSF ptau235 (Simoa) | 0.61 | 0.32-0.89 | n.s |
| Plasma Aβ42/40 (RocheNTK) | 0.64 | 0.48-0.80 | n.s |
| Plasma ptau231 (Simoa) | **0.74** | **0.53-0.97** | **0.009** |
| Plasma ptau181 (Simoa) | **0.77** | **0.56-0.98** | **0.003** |
| Plasma ptau217 (Lilly) | **0.77** | **0.59-0.96** | **0.003** |
| Plasma ptau181/Aβ42 (RocheNTK) | **0.79** | **0.65-0.93** | **0.002** |
| Plasma ptau181 (RocheNTK) | **0.79** | **0.65-0.93** | **0.002** |
| Plasma ptau181/Aβ40 (RocheNTK) | **0.80** | **0.66-0.94** | **0.002** |
| CSF NTAtau | **0.84** | **0.72-0.97** | **0.005** |
| CSF Aβ42/40 ratio | **0.88** | **0.77-0.99** | **<0.001** |
| Centiloid | **0.89** | **0.81-0.97** | **<0.0001** |
| CSF ptau181/Aβ40 | **0.91** | **0.84-0.98** | **<0.0001** |
| CSF ptau205 (Simoa) | **0.91** | **0.83-1.00** | **<0.0001** |
| CSF ptau181/Aβ42 | **0.91** | **0.82-1.00** | **<0.001** |
| CSF ptau217 (Lilly) | **0.96** | **0.91-1.00** | **<0.0001** |

Table_S4. Area under curve (AUC; 95%CI), corresponding p-value of core AD fluid biomarkers and Centiloid for predicting Braak I/II positivity, extracted from ROC analysis.

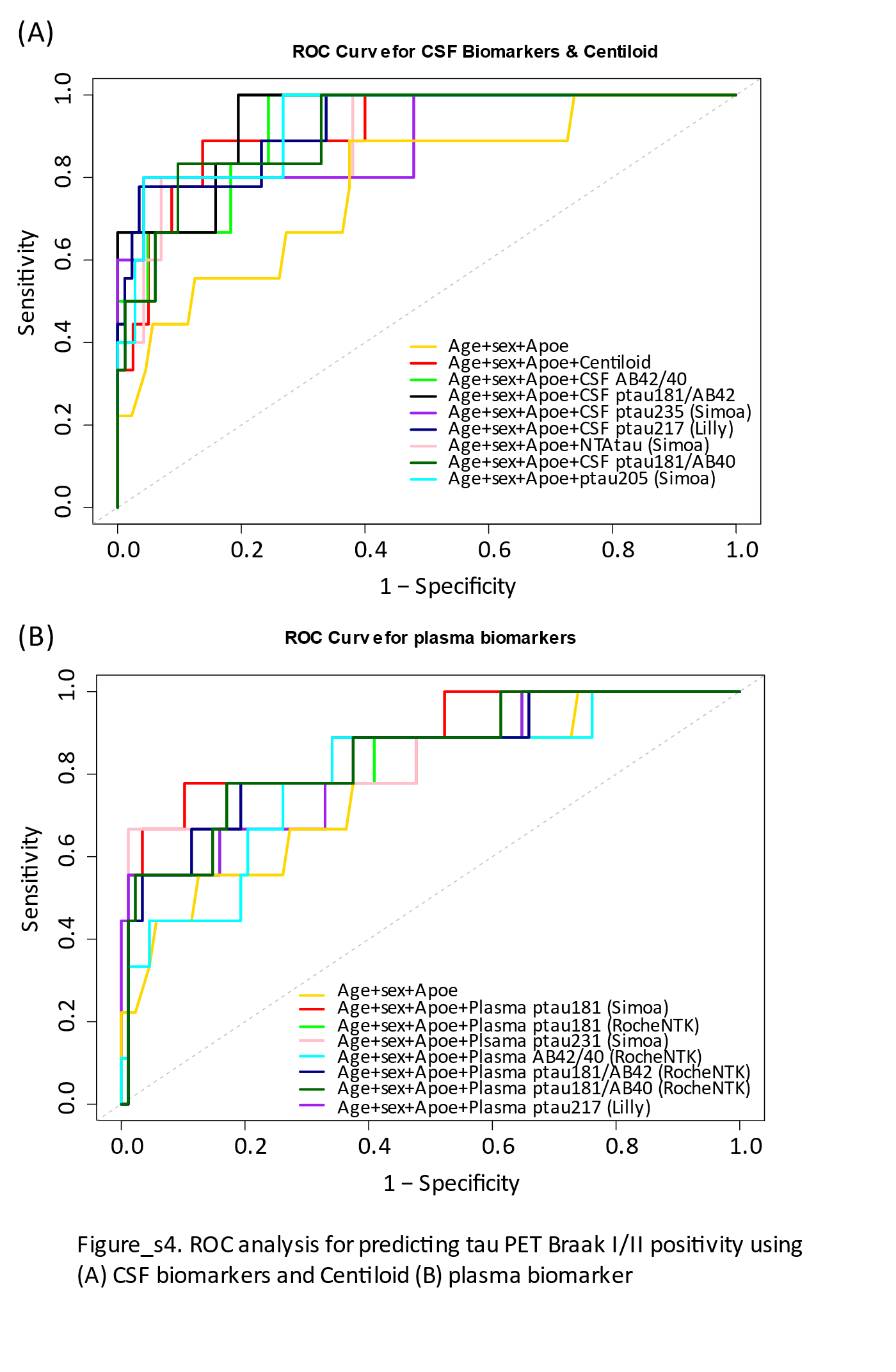

Figure_S6. ROC of the CSF (A) and plasma (B) biomarkers, adjusted for age, sex, and APOE ɛ4 carriership.

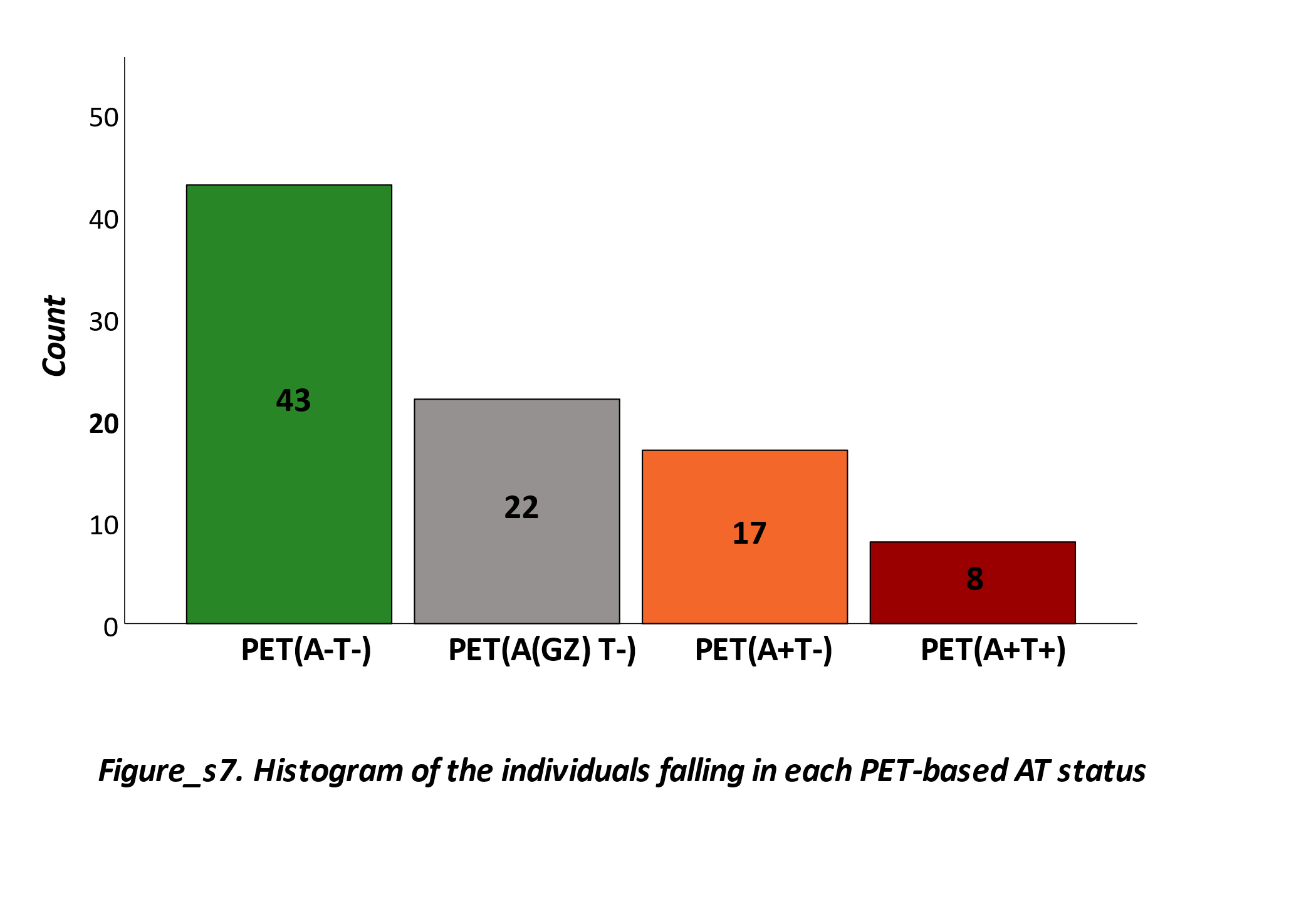

Figure_S7. Histogram of number of individuals in each PET-based stage

|  | *Total* | *A-T-* | *A(Gz)T-* | *A+T-* | *A+T+* |
| --- | --- | --- | --- | --- | --- |
| *N* | 99 | 42 | 21 | 17 | 9 |
| *Age, Mean (SD) [53-78 years]* | 65.15±5.00 | 63.42±4.18 | 65.38±5.15 | 67.06±4.95 | 69.66±4.58 |
| *Sex, N (%) [Female]* | 57 (57.57%) | 23 (54.76%) | 11 (52.38%) | 13 (76.47%) | 4 (44.44%) |
| *APOE-ε4, N (%) [Carriers]* | 63 (63.63%) | 24 (57.14%) | 13 (61.90%) | 13 (76.47%) | 6 (66.66%) |
| *Years of Education* | 13.35±3.78 | 14.28±4.12 | 13.05±3.95 | 13.48±4.27 | 13.00±2.23 |
| *CSF Aβ42/40, Mean (SD)* | 0.06±0.02 | 0.08±0.02 | 0.05±0.01 | 0.04±0.006 | 0.04±0.009 |
| *CSF ptau181, Mean (SD), pg/mL* | 21.92±9.40 | 16.46±7.15 | 18.69±7.08 | 24.05±6.33 | 33.28±9.03 |
| *CSF ptau181/Aβ40* | 0.001±0.00031 | 0.0009±0.0001 | 0.001±0.0002 | 0.001±0.0002 | 0.001±0.0003 |
| *CSF ptau181/Aβ42* | 0.02±0.013 | 0.01±0.006 | 0.02±0.01 | 0.03±0.008 | 0.04±0.01 |
| *Centiloid* | 22.51±26.25 | 0.68±7.80 | 23.06±8.50 | 57.44±14.92 | 57.24±16.52 |
| *CenTauR z-MetaROI* | 1.09±2.65 | 0.20±1.51 | 0.63±1.33 | 1.47±1.74 | 6.51±4.95 |
| *Hippocampal volume (mm^3^)* | 8860.14±1521.54 | 8806.12±991.28 | 8387.34±1050.71 | 8393.33±1002.85 | 8495.24±1833.92 |
| *Z-Scored PACC, Mean (SD)* | 0.05±0.62 | 0.23±0.58 | -0.05±0.62 | -0.05±0.63 | -0.36±0.63 |

Table_S5. Demographic information, cognition, Centiloid, and CSF biomarkers of the participants stratified by PET-based staging

| **Plasma Biomarker** | **Group1** | **Group2** | **n1** | **n2** | **statistic** | **p.adj.signif** |
| --- | --- | --- | --- | --- | --- | --- |
| **AB42/40 (RocheNTK)** | **A-T-** | **A(Gz)T-** | **42** | **21** | **197** | ******* |
| **AB42/40 (RocheNTK)** | **A-T-** | **A+T-** | **42** | **17** | **60** | ******** |
| **AB42/40 (RocheNTK)** | **A-T-** | **A+T+** | **42** | **9** | **67** | ****** |
| AB42/40 (RocheNTK) | A(Gz)T- | A+T- | 21 | 17 | 117 | ns |
| AB42/40 (RocheNTK) | A(Gz)T- | A+T+ | 21 | 9 | 92 | ns |
| AB42/40 (RocheNTK) | A+T- | A+T+ | 17 | 9 | 100 | ns |
| Ptau231 (Simoa) | A-T- | A(Gz)T- | 42 | 21 | 389 | ns |
| **Ptau231 (Simoa)** | **A-T-** | **A+T-** | **42** | **17** | **131** | ******* |
| **Ptau231 (Simoa)** | **A-T-** | **A+T+** | **42** | **9** | **72** | ****** |
| **Ptau231 (Simoa)** | **A(Gz)T-** | **A+T-** | **21** | **17** | **96** | ***** |
| Ptau231 (Simoa) | A(Gz)T- | A+T+ | 21 | 9 | 48 | ns |
| Ptau231 (Simoa) | A+T- | A+T+ | 17 | 9 | 62 | ns |
| **Ptau181/AB40 (RocheNTK)** | **A-T-** | **A(Gz)T-** | **42** | **21** | **192** | ******* |
| **Ptau181/AB40 (RocheNTK)** | **A-T-** | **A+T-** | **42** | **17** | **60** | ******** |
| **Ptau181/AB40 (RocheNTK)** | **A-T-** | **A+T+** | **42** | **9** | **28** | ******** |
| Ptau181/AB40 (RocheNTK) | A(Gz)T- | A+T- | 21 | 17 | 119 | ns |
| Ptau181/AB40 (RocheNTK) | A(Gz)T- | A+T+ | 21 | 9 | 51 | ns |
| Ptau181/AB40 (RocheNTK) | A+T- | A+T+ | 17 | 9 | 63 | ns |
| **Ptau181 (RocheNTK)** | **A-T-** | **A(Gz)T-** | **42** | **21** | **210** | ****** |
| **Ptau181 (RocheNTK)** | **A-T-** | **A+T-** | **42** | **17** | **47.5** | ******** |
| **Ptau181 (RocheNTK)** | **A-T-** | **A+T+** | **42** | **9** | **32** | ******* |
| **Ptau181 (RocheNTK)** | **A(Gz)T-** | **A+T-** | **21** | **17** | **83.5** | ***** |
| Ptau181 (RocheNTK) | A(Gz)T- | A+T+ | 21 | 9 | 48 | ns |
| Ptau181 (RocheNTK) | A+T- | A+T+ | 17 | 9 | 74 | ns |
| **Ptau217 (Lilly)** | **A-T-** | **A(Gz)T-** | **42** | **21** | **259** | ***** |
| **Ptau217 (Lilly)** | **A-T-** | **A+T-** | **42** | **17** | **82** | ******** |
| **Ptau217 (Lilly)** | **A-T-** | **A+T+** | **42** | **9** | **53** | ****** |
| **Ptau217 (Lilly)** | **A(Gz)T-** | **A+T-** | **21** | **17** | **71** | ****** |
| **Ptau217 (Lilly)** | **A(Gz)T-** | **A+T+** | **21** | **9** | **44** | ***** |
| Ptau217 (Lilly) | A+T- | A+T+ | 17 | 9 | 67 | ns |
| **Ptau181/AB42 (RocheNTK)** | **A-T-** | **A(Gz)T-** | **42** | **21** | **138** | ******** |
| **Ptau181/AB42 (RocheNTK)** | **A-T-** | **A+T-** | **42** | **17** | **27** | ******** |
| **Ptau181/AB42 (RocheNTK)** | **A-T-** | **A+T+** | **42** | **9** | **19** | ******** |
| Ptau181/AB42 (RocheNTK) | A(Gz)T- | A+T- | 21 | 17 | 112 | ns |
| Ptau181/AB42 (RocheNTK) | A(Gz)T- | A+T+ | 21 | 9 | 54 | ns |
| Ptau181/AB42 (RocheNTK) | A+T- | A+T+ | 17 | 9 | 70 | ns |
| Ptau181 (Simoa) | A-T- | A(Gz)T- | 42 | 21 | 308.5 | ns |
| **Ptau181 (Simoa)** | **A-T-** | **A+T-** | **42** | **17** | **103** | ******* |
| **Ptau181 (Simoa)** | **A-T-** | **A+T+** | **42** | **9** | **63.5** | ****** |
| **Ptau181 (Simoa)** | **A(Gz)T-** | **A+T-** | **21** | **17** | **90** | ***** |
| **Ptau181 (Simoa)** | **A(Gz)T-** | **A+T+** | **21** | **9** | **44** | ***** |
| Ptau181 (Simoa) | A+T- | A+T+ | 17 | 9 | 57 | ns |

Table_S6. Pairwise group comparisons of plasma biomarker levels across Amyloid and tau PET stages. The table summarizes statistical comparisons of plasma biomarker concentrations between 4 different groups defined by amyloid (A) and tau (T) PET status: A-T-, A(Gz)T-, A+T-, and A+T+. n1 and n2 refer to the sample sizes in each group. Significance is denoted as follows: p ≤ 0.05 (), p ≤ 0.01 (), p ≤ 0.001 (), and p ≤ 0.0001 (****); "ns" indicates non-significant results (p > 0.05). Stronger group differences are consistently observed for more advanced amyloid and tau pathology stages, particularly between A-T- and A+T+. All analysis done with R, and p-values are adjusted for FDR.

| **CSF Biomarkers and Centiloid** | **group1** | **group2** | **n1** | **n2** | **statistic** | **p.adj.signif** |
| --- | --- | --- | --- | --- | --- | --- |
| NTAtau (Simoa) | A-T- | A(Gz)T- | 34 | 16 | 213 | ns |
| **NTAtau (Simoa)** | **A-T-** | **A+T-** | **34** | **13** | **113** | ***** |
| **NTAtau (Simoa)** | **A-T-** | **A+T+** | **34** | **5** | **17** | ****** |
| NTAtau (Simoa) | A(Gz)T- | A+T- | 16 | 13 | 68 | ns |
| **NTAtau (Simoa)** | **A(Gz)T-** | **A+T+** | **16** | **5** | **10** | ***** |
| NTAtau (Simoa) | A+T- | A+T+ | 13 | 5 | 14 | ns |
| Ptau181 (RocheElecsys) | A-T- | A(Gz)T- | 41 | 17 | 281 | ns |
| **Ptau181 (RocheElecsys)** | **A-T-** | **A+T-** | **41** | **16** | **122** | ******* |
| **Ptau181 (RocheElecsys)** | **A-T-** | **A+T+** | **41** | **6** | **12** | ******* |
| **Ptau181 (RocheElecsys)** | **A(Gz)T-** | **A+T-** | **17** | **16** | **71** | ***** |
| **Ptau181 (RocheElecsys)** | **A(Gz)T-** | **A+T+** | **17** | **6** | **7** | ****** |
| Ptau181 (RocheElecsys) | A+T- | A+T+ | 16 | 6 | 24 | ns |
| Ptau181 (RocheNTK) | A-T- | A(Gz)T- | 41 | 17 | 282 | ns |
| **Ptau181 (RocheNTK)** | **A-T-** | **A+T-** | **41** | **16** | **124** | ****** |
| **Ptau181 (RocheNTK)** | **A-T-** | **A+T+** | **41** | **6** | **10** | ****** |
| **Ptau181 (RocheNTK)** | **A(Gz)T-** | **A+T-** | **17** | **16** | **69** | ***** |
| **Ptau181 (RocheNTK)** | **A(Gz)T-** | **A+T+** | **17** | **6** | **7** | ****** |
| **Ptau181 (RocheNTK)** | **A+T-** | **A+T+** | **16** | **6** | **19** | ***** |
| Ptau205 (Simoa) | A-T- | A(Gz)T- | 34 | 16 | 192 | ns |
| **Ptau205 (Simoa)** | **A-T-** | **A+T-** | **34** | **13** | **67** | ******* |
| **Ptau205 (Simoa)** | **A-T-** | **A+T+** | **34** | **5** | **4** | ******* |
| Ptau205 (Simoa) | A(Gz)T- | A+T- | 16 | 13 | 58 | ns |
| **Ptau205 (Simoa)** | **A(Gz)T-** | **A+T+** | **16** | **5** | **5** | ****** |
| Ptau205 (Simoa) | A+T- | A+T+ | 13 | 5 | 18 | ns |
| Ptau235 (Simoa) | A-T- | A(Gz)T- | 34 | 16 | 172 | ns |
| **Ptau235 (Simoa)** | **A-T-** | **A+T-** | **34** | **13** | **65** | ******* |
| **Ptau235 (Simoa)** | **A-T-** | **A+T+** | **34** | **5** | **6** | ******* |
| Ptau235 (Simoa) | A(Gz)T- | A+T- | 16 | 13 | 58 | ns |
| **Ptau235 (Simoa)** | **A(Gz)T-** | **A+T+** | **16** | **5** | **10** | ***** |
| Ptau235 (Simoa) | A+T- | A+T+ | 13 | 5 | 19 | ns |
| **Ptau217 (Lilly)** | **A-T-** | **A(Gz)T-** | **41** | **17** | **180** | ****** |
| **Ptau217 (Lilly)** | **A-T-** | **A+T-** | **41** | **16** | **34** | ******** |
| **Ptau217 (Lilly)** | **A-T-** | **A+T+** | **41** | **6** | **2** | ******** |
| **Ptau217 (Lilly)** | **A(Gz)T-** | **A+T-** | **17** | **16** | **51** | ****** |
| **Ptau217 (Lilly)** | **A(Gz)T-** | **A+T+** | **17** | **6** | **3** | ******* |
| **Ptau217 (Lilly)** | **A+T-** | **A+T+** | **16** | **6** | **11** | ****** |
| **AB42/40 (RocheNTK)** | **A-T-** | **A(Gz)T-** | **41** | **17** | **100** | ******** |
| **AB42/40 (RocheNTK)** | **A-T-** | **A+T-** | **41** | **16** | **23** | ******** |
| **AB42/40 (RocheNTK)** | **A-T-** | **A+T+** | **41** | **6** | **5** | ******** |
| AB42/40 (RocheNTK) | A(Gz)T- | A+T- | 17 | 16 | 83 | ns |
| **AB42/40 (RocheNTK)** | **A(Gz)T-** | **A+T+** | **17** | **6** | **17** | ***** |
| AB42/40 (RocheNTK) | A+T- | A+T+ | 16 | 6 | 27 | ns |
| **Ptau181/AB40 (RocheNTK)** | **A-T-** | **A(Gz)T-** | **41** | **17** | **143** | ******* |
| **Ptau181/AB40 (RocheNTK)** | **A-T-** | **A+T-** | **41** | **16** | **35** | ******** |
| **Ptau181/AB40 (RocheNTK)** | **A-T-** | **A+T+** | **41** | **6** | **4** | ******** |
| **Ptau181/AB40 (RocheNTK)** | **A(Gz)T-** | **A+T-** | **17** | **16** | **70** | ***** |
| **Ptau181/AB40 (RocheNTK)** | **A(Gz)T-** | **A+T+** | **17** | **6** | **10** | ****** |
| **Ptau181/AB40 (RocheNTK)** | **A+T-** | **A+T+** | **16** | **6** | **17** | ***** |
| **Centiloid** | **A-T-** | **A(Gz)T-** | **42** | **21** | **0** | ******** |
| **Centiloid** | **A-T-** | **A+T-** | **42** | **17** | **0** | ******** |
| **Centiloid** | **A-T-** | **A+T+** | **42** | **9** | **0** | ******** |
| **Centiloid** | **A(Gz)T-** | **A+T-** | **21** | **17** | **0** | ******** |
| **Centiloid** | **A(Gz)T-** | **A+T+** | **21** | **9** | **10** | ******** |
| Centiloid | A+T- | A+T+ | 17 | 9 | 69 | ns |
| **Ptau181/AB42 (RocheNTK)** | **A-T-** | **A(Gz)T-** | **41** | **17** | **83** | ******** |
| **Ptau181/AB42 (RocheNTK)** | **A-T-** | **A+T-** | **41** | **16** | **17** | ******** |
| **Ptau181/AB42 (RocheNTK)** | **A-T-** | **A+T+** | **41** | **6** | **3** | ******** |
| **Ptau181/AB42 (RocheNTK)** | **A(Gz)T-** | **A+T-** | **17** | **16** | **71** | ***** |
| **Ptau181/AB42 (RocheNTK)** | **A(Gz)T-** | **A+T+** | **17** | **6** | **9** | ****** |
| **Ptau181/AB42 (RocheNTK)** | **A+T-** | **A+T+** | **16** | **6** | **20** | ***** |

Table_S7. Pairwise group comparisons of CSF biomarkers levels and Centiloid across Amyloid and tau PET stages. The table summarizes statistical comparisons of CSF biomarker concentrations and Centiloid values between 4 different groups defined by amyloid (A) and tau (T) PET status: A-T-, A(Gz)T-, A+T-, and A+T+. n1 and n2 refer to the sample sizes in each group. Significance is denoted as follows: p ≤ 0.05 (), p ≤ 0.01 (), p ≤ 0.001 (), and p ≤ 0.0001 (****); "ns" indicates non-significant results (p > 0.05). Stronger group differences are consistently observed for more advanced amyloid and tau pathology stages, particularly between A-T- and A+T+. All analysis done with R, and p-values are adjusted for FDR

| **Cognition** | **group1** | **group2** | **n1** | **n2** | **statistic** | **p.adj.signif** |
| --- | --- | --- | --- | --- | --- | --- |
| Attention | A-T- | A(Gz)T- | 42 | 21 | 543 | ns |
| Attention | A-T- | A+T- | 42 | 17 | 509 | ns |
| Attention | A-T- | A+T+ | 42 | 9 | 243 | ns |
| Attention | A(Gz)T- | A+T- | 21 | 17 | 214 | ns |
| Attention | A(Gz)T- | A+T+ | 21 | 9 | 99 | ns |
| Attention | A+T- | A+T+ | 17 | 9 | 67 | ns |
| Memory | A-T- | A(Gz)T- | 42 | 21 | 486 | ns |
| Memory | A-T- | A+T- | 42 | 16 | 367 | ns |
| Memory | A-T- | A+T+ | 42 | 8 | 223 | ns |
| Memory | A(Gz)T- | A+T- | 21 | 16 | 165 | ns |
| Memory | A(Gz)T- | A+T+ | 21 | 8 | 106 | ns |
| Memory | A+T- | A+T+ | 16 | 8 | 81 | ns |
| Executive | A-T- | A(Gz)T- | 42 | 21 | 514 | ns |
| **Executive** | **A-T-** | **A+T-** | **42** | **17** | **546** | ***** |
| Executive | A-T- | A+T+ | 42 | 8 | 204 | ns |
| Executive | A(Gz)T- | A+T- | 21 | 17 | 246 | ns |
| Executive | A(Gz)T- | A+T+ | 21 | 8 | 90 | ns |
| Executive | A+T- | A+T+ | 17 | 8 | 47 | ns |
| Language | A-T- | A(Gz)T- | 42 | 21 | 543.5 | ns |
| Language | A-T- | A+T- | 42 | 16 | 383 | ns |
| Language | A-T- | A+T+ | 42 | 8 | 222.5 | ns |
| Language | A(Gz)T- | A+T- | 21 | 16 | 155.5 | ns |
| Language | A(Gz)T- | A+T+ | 21 | 8 | 91 | ns |
| Language | A+T- | A+T+ | 16 | 8 | 79 | ns |
| Visual | A-T- | A(Gz)T- | 42 | 21 | 518 | ns |
| Visual | A-T- | A+T- | 42 | 17 | 454.5 | ns |
| Visual | A-T- | A+T+ | 42 | 9 | 282.5 | ns |
| Visual | A(Gz)T- | A+T- | 21 | 17 | 195 | ns |
| Visual | A(Gz)T- | A+T+ | 21 | 9 | 122.5 | ns |
| Visual | A+T- | A+T+ | 17 | 9 | 91 | ns |
| PACC | A-T- | A(Gz)T- | 42 | 21 | 518 | ns |
| PACC | A-T- | A+T- | 42 | 17 | 454.5 | ns |
| PACC | A-T- | A+T+ | 42 | 9 | 282.5 | ns |
| PACC | A(Gz)T- | A+T- | 21 | 17 | 195 | ns |
| PACC | A(Gz)T- | A+T+ | 21 | 9 | 122.5 | ns |
| PACC | A+T- | A+T+ | 17 | 9 | 91 | ns |

Table_S8. Pairwise group comparisons of attention, memory, executive function, language, visual, and PACC levels across Amyloid and tau PET stages. The table summarizes statistical comparisons of cognitive scores between 4 different groups defined by amyloid (A) and tau (T) PET status: A-T-, A(Gz)T-, A+T-, and A+T+. n1 and n2 refer to the sample sizes in each group. Significance is denoted as follows: p ≤ 0.05 (), p ≤ 0.01 (), p ≤ 0.001 (), and p ≤ 0.0001 (****); "ns" indicates non-significant results (p > 0.05). All analysis done with R, and p-values are adjusted for FDR.

| **Plasma Biomarker** | **Group1** | **Group2** | **n1** | **n2** | **statistic** | **p.adj.signif** |
| --- | --- | --- | --- | --- | --- | --- |
| **AB42/40 (RocheNTK)** | **A-T-** | **A(Gz)T-** | **42** | **21** | **685** | ******* |
| **AB42/40 (RocheNTK)** | **A-T-** | **A+T-** | **42** | **17** | **654** | ******** |
| **AB42/40 (RocheNTK)** | **A-T-** | **A+T+** | **42** | **9** | **311** | ****** |
| AB42/40 (RocheNTK) | A(Gz)T- | A+T- | 21 | 17 | 240 | ns |
| AB42/40 (RocheNTK) | A(Gz)T- | A+T+ | 21 | 9 | 97 | ns |
| AB42/40 (RocheNTK) | A+T- | A+T+ | 17 | 9 | 53 | ns |
| Ptau231 (Simoa) | A-T- | A(Gz)T- | 42 | 21 | 389 | ns |
| **Ptau231 (Simoa)** | **A-T-** | **A+T-** | **42** | **17** | **131** | ******* |
| **Ptau231 (Simoa)** | **A-T-** | **A+T+** | **42** | **9** | **72** | ****** |
| **Ptau231 (Simoa)** | **A(Gz)T-** | **A+T-** | **21** | **17** | **96** | ***** |
| Ptau231 (Simoa) | A(Gz)T- | A+T+ | 21 | 9 | 48 | ns |
| Ptau231 (Simoa) | A+T- | A+T+ | 17 | 9 | 62 | ns |
| **Ptau181/AB40 (RocheNTK)** | **A-T-** | **A(Gz)T-** | **42** | **21** | **192** | ******* |
| **Ptau181/AB40 (RocheNTK)** | **A-T-** | **A+T-** | **42** | **17** | **60** | ******** |
| **Ptau181/AB40 (RocheNTK)** | **A-T-** | **A+T+** | **42** | **9** | **28** | ******** |
| Ptau181/AB40 (RocheNTK) | A(Gz)T- | A+T- | 21 | 17 | 119 | ns |
| Ptau181/AB40 (RocheNTK) | A(Gz)T- | A+T+ | 21 | 9 | 51 | ns |
| Ptau181/AB40 (RocheNTK) | A+T- | A+T+ | 17 | 9 | 63 | ns |
| **Ptau181 (RocheNTK)** | **A-T-** | **A(Gz)T-** | **42** | **21** | **210** | ****** |
| **Ptau181 (RocheNTK)** | **A-T-** | **A+T-** | **42** | **17** | **47.5** | ******** |
| **Ptau181 (RocheNTK)** | **A-T-** | **A+T+** | **42** | **9** | **32** | ******* |
| **Ptau181 (RocheNTK)** | **A(Gz)T-** | **A+T-** | **21** | **17** | **83.5** | ***** |
| Ptau181 (RocheNTK) | A(Gz)T- | A+T+ | 21 | 9 | 48 | ns |
| Ptau181 (RocheNTK) | A+T- | A+T+ | 17 | 9 | 74 | ns |
| **Ptau217 (Lilly)** | **A-T-** | **A(Gz)T-** | **42** | **21** | **259** | ***** |
| **Ptau217 (Lilly)** | **A-T-** | **A+T-** | **42** | **17** | **82** | ******** |
| **Ptau217 (Lilly)** | **A-T-** | **A+T+** | **42** | **9** | **53** | ****** |
| **Ptau217 (Lilly)** | **A(Gz)T-** | **A+T-** | **21** | **17** | **71** | ****** |
| **Ptau217 (Lilly)** | **A(Gz)T-** | **A+T+** | **21** | **9** | **44** | ***** |
| Ptau217 (Lilly) | A+T- | A+T+ | 17 | 9 | 67 | ns |
| **Ptau181/AB42 (RocheNTK)** | **A-T-** | **A(Gz)T-** | **42** | **21** | **138** | ******** |
| **Ptau181/AB42 (RocheNTK)** | **A-T-** | **A+T-** | **42** | **17** | **27** | ******** |
| **Ptau181/AB42 (RocheNTK)** | **A-T-** | **A+T+** | **42** | **9** | **19** | ******** |
| Ptau181/AB42 (RocheNTK) | A(Gz)T- | A+T- | 21 | 17 | 112 | ns |
| Ptau181/AB42 (RocheNTK) | A(Gz)T- | A+T+ | 21 | 9 | 54 | ns |
| Ptau181/AB42 (RocheNTK) | A+T- | A+T+ | 17 | 9 | 70 | ns |
| Ptau181 (Simoa) | A-T- | A(Gz)T- | 42 | 21 | 308.5 | ns |
| **Ptau181 (Simoa)** | **A-T-** | **A+T-** | **42** | **17** | **103** | ******* |
| **Ptau181 (Simoa)** | **A-T-** | **A+T+** | **42** | **9** | **63.5** | ****** |
| **Ptau181 (Simoa)** | **A(Gz)T-** | **A+T-** | **21** | **17** | **90** | ***** |
| **Ptau181 (Simoa)** | **A(Gz)T-** | **A+T+** | **21** | **9** | **44** | ***** |
| Ptau181 (Simoa) | A+T- | A+T+ | 17 | 9 | 57 | ns |

Table_S9. Pairwise group comparisons of the fold of changes for plasma biomarkers across Amyloid and tau PET stages. The table summarizes statistical comparisons of the fold of changes in plasma biomarker concentrations between 4 different groups defined by amyloid (A) and tau (T) PET status: A-T-, A(Gz)T-, A+T-, and A+T+. n1 and n2 refer to the sample sizes in each group. Significance is denoted as follows: p ≤ 0.05 (), p ≤ 0.01 (), p ≤ 0.001 (), and p ≤ 0.0001 (****); "ns" indicates non-significant results (p > 0.05). All analysis done with R, and p-values are adjusted for FDR.

| **Biomarker** | **group1** | **group2** | **n1** | **n2** | **statistic** | **p.adj.signif** |
| --- | --- | --- | --- | --- | --- | --- |
| NTAtau (Simoa) | A-T- | A(Gz)T- | 34 | 16 | 213 | ns |
| **NTAtau (Simoa)** | **A-T-** | **A+T-** | **34** | **13** | **113** | ***** |
| **NTAtau (Simoa)** | **A-T-** | **A+T+** | **34** | **5** | **17** | ****** |
| NTAtau (Simoa) | A(Gz)T- | A+T- | 16 | 13 | 68 | ns |
| **NTAtau (Simoa)** | **A(Gz)T-** | **A+T+** | **16** | **5** | **10** | ***** |
| NTAtau (Simoa) | A+T- | A+T+ | 13 | 5 | 14 | ns |
| Ptau181 (RocheElecsys) | A-T- | A(Gz)T- | 41 | 17 | 281 | ns |
| **Ptau181 (RocheElecsys)** | **A-T-** | **A+T-** | **41** | **16** | **122** | ******* |
| **Ptau181 (RocheElecsys)** | **A-T-** | **A+T+** | **41** | **6** | **12** | ******* |
| **Ptau181 (RocheElecsys)** | **A(Gz)T-** | **A+T-** | **17** | **16** | **71** | ***** |
| **Ptau181 (RocheElecsys)** | **A(Gz)T-** | **A+T+** | **17** | **6** | **7** | ****** |
| Ptau181 (RocheElecsys) | A+T- | A+T+ | 16 | 6 | 24 | ns |
| Ptau181 (RocheNTK) | A-T- | A(Gz)T- | 41 | 17 | 282 | ns |
| **Ptau181 (RocheNTK)** | **A-T-** | **A+T-** | **41** | **16** | **124** | ****** |
| **Ptau181 (RocheNTK)** | **A-T-** | **A+T+** | **41** | **6** | **10** | ****** |
| **Ptau181 (RocheNTK)** | **A(Gz)T-** | **A+T-** | **17** | **16** | **69** | ***** |
| **Ptau181 (RocheNTK)** | **A(Gz)T-** | **A+T+** | **17** | **6** | **7** | ****** |
| **Ptau181 (RocheNTK)** | **A+T-** | **A+T+** | **16** | **6** | **19** | ***** |
| Ptau205 (Simoa) | A-T- | A(Gz)T- | 34 | 16 | 192 | ns |
| **Ptau205 (Simoa)** | **A-T-** | **A+T-** | **34** | **13** | **67** | ******* |
| **Ptau205 (Simoa)** | **A-T-** | **A+T+** | **34** | **5** | **4** | ******* |
| Ptau205 (Simoa) | A(Gz)T- | A+T- | 16 | 13 | 58 | ns |
| **Ptau205 (Simoa)** | **A(Gz)T-** | **A+T+** | **16** | **5** | **5** | ****** |
| Ptau205 (Simoa) | A+T- | A+T+ | 13 | 5 | 18 | ns |
| Ptau235 (Simoa) | A-T- | A(Gz)T- | 34 | 16 | 172 | ns |
| **Ptau235 (Simoa)** | **A-T-** | **A+T-** | **34** | **13** | **65** | ******* |
| **Ptau235 (Simoa)** | **A-T-** | **A+T+** | **34** | **5** | **6** | ******* |
| Ptau235 (Simoa) | A(Gz)T- | A+T- | 16 | 13 | 58 | ns |
| **Ptau235 (Simoa)** | **A(Gz)T-** | **A+T+** | **16** | **5** | **10** | ***** |
| Ptau235 (Simoa) | A+T- | A+T+ | 13 | 5 | 19 | ns |
| **Ptau217 (Lilly)** | **A-T-** | **A(Gz)T-** | **41** | **17** | **180** | ****** |
| **Ptau217 (Lilly)** | **A-T-** | **A+T-** | **41** | **16** | **34** | ******** |
| **Ptau217 (Lilly)** | **A-T-** | **A+T+** | **41** | **6** | **2** | ******** |
| **Ptau217 (Lilly)** | **A(Gz)T-** | **A+T-** | **17** | **16** | **51** | ****** |
| **Ptau217 (Lilly)** | **A(Gz)T-** | **A+T+** | **17** | **6** | **3** | ******* |
| **Ptau217 (Lilly)** | **A+T-** | **A+T+** | **16** | **6** | **11** | ****** |
| **AB42/40 (RocheNTK)** | **A-T-** | **A(Gz)T-** | **41** | **17** | **597** | ******** |
| **AB42/40 (RocheNTK)** | **A-T-** | **A+T-** | **41** | **16** | **633** | ******** |
| **AB42/40 (RocheNTK)** | **A-T-** | **A+T+** | **41** | **6** | **241** | ******** |
| AB42/40 (RocheNTK) | A(Gz)T- | A+T- | 17 | 16 | 189 | ns |
| **AB42/40 (RocheNTK)** | **A(Gz)T-** | **A+T+** | **17** | **6** | **85** | ***** |
| AB42/40 (RocheNTK) | A+T- | A+T+ | 16 | 6 | 69 | ns |
| **Ptau181/AB40 (RocheNTK)** | **A-T-** | **A(Gz)T-** | **41** | **17** | **143** | ******* |
| **Ptau181/AB40 (RocheNTK)** | **A-T-** | **A+T-** | **41** | **16** | **35** | ******** |
| **Ptau181/AB40 (RocheNTK)** | **A-T-** | **A+T+** | **41** | **6** | **4** | ******** |
| **Ptau181/AB40 (RocheNTK)** | **A(Gz)T-** | **A+T-** | **17** | **16** | **70** | ***** |
| **Ptau181/AB40 (RocheNTK)** | **A(Gz)T-** | **A+T+** | **17** | **6** | **10** | ****** |
| **Ptau181/AB40 (RocheNTK)** | **A+T-** | **A+T+** | **16** | **6** | **17** | ***** |
| **Ptau181/AB42 (RocheNTK)** | **A-T-** | **A(Gz)T-** | **41** | **17** | **83** | ******** |
| **Ptau181/AB42 (RocheNTK)** | **A-T-** | **A+T-** | **41** | **16** | **17** | ******** |
| **Ptau181/AB42 (RocheNTK)** | **A-T-** | **A+T+** | **41** | **6** | **3** | ******** |
| **Ptau181/AB42 (RocheNTK)** | **A(Gz)T-** | **A+T-** | **17** | **16** | **71** | ***** |
| **Ptau181/AB42 (RocheNTK)** | **A(Gz)T-** | **A+T+** | **17** | **6** | **9** | ****** |
| **Ptau181/AB42 (RocheNTK)** | **A+T-** | **A+T+** | **16** | **6** | **20** | ***** |

Table_S10. Pairwise group comparisons of the fold of changes for CSF biomarkers across Amyloid and tau PET stages. The table summarizes statistical comparisons of the fold of changes in CSF biomarker concentrations between 4 different groups defined by amyloid (A) and tau (T) PET status: A-T-, A(Gz)T-, A+T-, and A+T+. n1 and n2 refer to the sample sizes in each group. Significance is denoted as follows: p ≤ 0.05 (), p ≤ 0.01 (), p ≤ 0.001 (), and p ≤ 0.0001 (****); "ns" indicates non-significant results (p > 0.05). All analysis done with R, and p-values are adjusted for FDR.
